# Clinical Correlates of Suicidal Ideation in Adults With Obsessive-Compulsive Disorder: An Interpretable Machine Learning Study

**DOI:** 10.64898/2026.05.31.26354549

**Authors:** Brian A. Zaboski, Elizabeth F. Mattera, Christopher Pittenger

## Abstract

**Introduction:** Depression accounts for much of the association between obsessive-compulsive disorder and suicidal ideation in clinical samples, but whether other features carry independent information is unclear. We examined what non-depression features reveal about suicidal ideation.

**Methods:** Adults with obsessive-compulsive disorder (*N* = 231) completed measures of quality of life, anxiety, physical symptoms, obsessive-compulsive symptoms and traits, and personality. Elastic net, random forest, and explainable boosting machines identified suicidal ideation (Beck Depression Inventory-II item 9) without depression measures in nested cross-validation. Secondary analyses compared these features with depression severity.

**Results:** Sixty-six participants (28.6%) endorsed suicidal ideation; 61 of these (92.4%) reported thoughts without intent to act. All models showed similar discrimination (area under the curve = .77–.78), as did depression severity alone (.77). Adding the features to depression severity yielded a small estimated gain (difference = .02, 95% confidence interval: −.03 to .06), and flexible models offered no reliable advantage. Most features, including trait anxiety and obsessive-compulsive symptom severity, were associated with suicidal ideation on their own, but these associations attenuated under mutual adjustment. Lower quality of life remained the clearest correlate.

**Conclusion:** Non-depression features identified suicidal ideation about as well as depression severity, consistent with substantial shared, distress-related information. Clinicians should ask about suicidal ideation directly, whatever a patient’s depressive symptoms, and consider overall distress in case formulation.

---

Obsessive-compulsive disorder (OCD) carries a substantially elevated risk of suicide. In a Swedish register study of 36,788 patients, the risk of death by suicide was roughly ten times that of matched controls, and 43% of those who died by suicide had no recorded psychiatric comorbidity (Fernández de la Cruz et al., 2017). The absence of a recorded diagnosis is not the absence of depressive symptoms, however, and in clinical samples depressive symptoms are among the strongest correlates of suicidal ideation (SI). In several clinical analyses, the association between OCD severity and SI weakened once depressive symptoms were taken into account—although findings vary with the outcome, symptom dimensions, and design (Angelakis et al., 2015).

These findings leave open what clinical features other than depression reveal about SI in OCD, and how much of that information overlaps with what depression severity already captures. Lower quality of life, anxiety, and particular obsessive-compulsive symptom dimensions have been linked to suicidality in OCD (Storch et al., 2017; Velloso et al., 2016), but these features share variance with depression. Earlier analyses of overlapping participants from our clinic found that anhedonia’s association with OCD severity disappeared once depression was controlled (Zaboski, Jones, et al., 2026) and that patients were better described by a single continuous dimension of general distress than by discrete subgroups (Zaboski, Jones, Mattera, et al., 2025), suggesting that overlap may be the central issue. The present study therefore examined how these clinical domains jointly characterize SI, excluding depression measures from the primary feature set so that the features could be compared directly with a depression benchmark.

Our goals are framed by two models. The tripartite model holds that anxiety and depression share a general negative-affect component, with low positive affect more specific to depression, and physiological hyperarousal more specific to anxiety (Clark & Watson, 1991). Relatedly, the integrated motivational–volitional (IMV) model locates the emergence of ideation in experiences of defeat and entrapment, which the burdens of OCD could foster, and separates that motivational phase from the volitional factors that govern progression to suicidal behavior (O’Connor & Kirtley, 2018). The present measures do not assess defeat or entrapment, so the IMV model provides context rather than a hypothesis under test.

A further question is whether these domains combine in ways that additive models miss. Systems perspectives hold that psychiatric features interact and that problems can emerge once stress exceeds a system’s capacity to absorb it (Borsboom, 2017; Kendler et al., 2011), and allostatic accounts attribute a cumulative physical toll to sustained stress (McEwen, 1998).

Neither specifies which statistical interactions to expect among questionnaire totals in a cross-sectional sample, so predictions drawn from them are theory-informed exploratory expectations. Explainable boosting machines (EBMs) can examine them while remaining interpretable, estimating a separate shape function for each feature, with optional pairwise interactions, rather than assuming constant, additive associations (Higgins et al., 2024; Nori et al., 2019).

We compared three algorithms, elastic-net logistic regression, random forest, and an EBM, and used the EBM for interpretation, because interpretable models are preferable to black-box alternatives when their performance is comparable (Rudin, 2019). As stated in a preprint of this study (Zaboski, Mattera, et al., 2026), we hypothesized that associations between obsessive-compulsive symptom burden and SI would be nonlinear, with quality of life and trait conscientiousness (grouped in the preprint as functional capacities) showing nonlinear protective associations. We further hypothesized that elevated SI would be characterized by interactive rather than additive phenotypes, such as severe obsessive-compulsive traits combined with elevated somatic symptom burden, a pairing derived from allostatic accounts for which a physical symptom checklist is only a loose proxy. Secondary analyses asked how to interpret the resulting profile in relation to depression: whether the features add information beyond depression severity or largely overlap with it, and whether any overlap reflects items that duplicate depressive symptoms.

## Methods

### Participants

All procedures were approved by the Yale University Institutional Review Board, and participants provided written informed consent. Data were drawn from an ongoing clinical-research registry at the Yale OCD Research Clinic, which has enrolled adults in a data repository since 2020; data for this study were extracted in May 2026. Participants were recruited through local and online advertisements and referrals from community providers, completed a telephone screen, and attended an intake evaluation. Self-report measures were collected in REDCap (Harris et al., 2009, 2019), and participants received $40 for completing them.

Adults aged 18–70 years with a primary or co-primary diagnosis of OCD were eligible. Diagnoses were established through structured and unstructured clinical assessment, using the Mini-International Neuropsychiatric Interview (Sheehan et al., 1998) or the Diagnostic Interview for Anxiety, Mood, and OCD and Related Neuropsychiatric Disorders (Tolin et al., 2018), and were confirmed by a board-certified psychologist or psychiatrist. Comorbid conditions were permitted when OCD was primary or co-primary. Exclusion criteria were a history of psychosurgery; current mania or psychosis; medical conditions affecting the central nervous system, medical instability, or traumatic brain injury with loss of consciousness exceeding 30 minutes; substance use disorders within the past 6 months other than mild tobacco, alcohol, or cannabis use disorder; and active suicidal ideation with a plan or intent. History of bipolar I disorder and autism spectrum disorder were also exclusionary. Of 236 registry records with questionnaire and demographic data, 5 lacked a response to the suicidal ideation item, leaving an analytic sample of 231. The sample size was set by registry availability rather than by a power calculation; with 66 participants endorsing SI and 15 features (4.4 events per feature), estimates for flexible models and individual features were expected to be imprecise.

### Measures

#### Suicidal ideation

The outcome was item 9 of the Beck Depression Inventory–II (BDI-II; (Beck et al., 1996), which asks about suicidal thoughts or wishes over the past 2 weeks on a 0–3 scale (0 = no thoughts of killing oneself; 1 = such thoughts without intending to act on them; 2 = wanting to kill oneself; 3 = would kill oneself given the chance). Responses were dichotomized as absent (0) or present (≥ 1). Because the item asks only about thoughts of killing oneself, it cannot distinguish SI from ego-dystonic intrusive thoughts about suicide, which are common in OCD (Mattera et al., 2024, 2025).

#### Depression severity (secondary analyse

Depression severity was indexed by the BDI-II total excluding item 9 (20 items; α = .92), which removes the outcome item from the covariate (cf. Zaboski, Jones, Mattera, et al., 2025). The Patient-Reported Outcomes Measurement Information System (PROMIS) Depression bank v1.0 T-score (Pilkonis et al., 2011), administered as a computerized adaptive test, served as a second depression measure in a sensitivity analysis. Because the bank includes an item about having no reason for living, it is not entirely free of suicide-related content.

#### Quality of life

The Quality of Life Enjoyment and Satisfaction Questionnaire–Short Form (Q-LES-Q-SF; Endicott et al., 1993) rates satisfaction across life domains over the past week from 1 (*very poor*) to 5 (*very good*). Following standard scoring, items 1–14 were summed (range 14–70; α = .87). Item 15, which is coded 0 when a participant is not taking medication, was used to derive current medication status (any medication vs. none).

#### Physical symptoms

Physical symptom distress was assessed with a 23-item checklist adapted from the Physical Symptom Distress subscale of the Rotterdam Symptom Checklist (de Haes et al., 2012). Participants rated how bothersome each symptom (e.g., headache, nausea, dry mouth) had been over the past week from 1 (*none*) to 4 (*severe*); totals range from 23 to 92 (α = .83).

#### Obsessive-compulsive symptoms and traits

The Dimensional Obsessive-Compulsive Scale (DOCS; Abramowitz et al., 2010) assesses the severity of contamination, responsibility for harm, unacceptable thoughts, and symmetry symptoms (20 items rated 0–4; total 0–80; α = .89). The Obsessive-Compulsive Trait Core Dimensions Questionnaire (OCTCDQ; Summerfeldt et al., 2001, 2014) assesses harm avoidance and incompleteness (10 items each, rated 1–5; subscale range 10–50; α = .91 for both). Only the subscales were analyzed, because the total score is their sum.

#### Personality

The Big Five Aspects Scale (BFAS; DeYoung et al., 2007) measures five domains, each comprising two aspects, with 100 items rated 1–5. Domain scores were item means (α = .79–.90). The two conscientiousness aspects, industriousness and orderliness, were used in a secondary analysis.

#### Anxiety

The State-Trait Anxiety Inventory (STAI; Spielberger et al., 1983) assesses state and trait anxiety (20 items each; range 20–80; α = .94 and .92, respectively). State-scale item 4 had been stored with reversed response coding in the electronic record and was recoded before scoring.

#### Demographic variables

Age and sex were included as features. Sex was coded as female versus other responses, with “prefer not to say” treated as missing. Gender identity was assessed separately and is reported descriptively.

### Analytic Plan

#### Scoring and missing data

Scale scores were computed from items. When at least 80% of a scale’s items were answered, scores were prorated from the available items; otherwise they were set to missing. Proration affected 14 of 2,772 scale scores (0.5%), and one participant’s OCTCDQ subscale scores were set to missing. Missingness on the features ranged from 0.4% to 6.9%. Missing values were imputed by *k*-nearest neighbors (*k* = 5) on standardized features; the imputation was fit to the training data within each cross-validation fold and applied to the corresponding test fold.

#### Features and models

The primary feature set comprised 15 variables: age, sex, medication status, quality of life, physical symptoms, DOCS total, OCTCDQ harm avoidance and incompleteness, the five BFAS domains, and state and trait anxiety. Depression measures were intentionally excluded from this set. Three algorithms were compared: elastic-net logistic regression, random forest, and an EBM (Nori et al., 2019), which was fit both with main effects only and with pairwise interactions. Hyperparameters were tuned over prespecified grids (Supplementary Table S1). Classes were not reweighted.

#### Validation

Performance was estimated with nested cross-validation (Cawley & Talbot, 2010; Vabalas et al., 2019) with an outer loop of five folds repeated five times (25 test folds) and an inner three-fold loop for tuning, with the area under the precision-recall curve as the tuning criterion. We report the areas under the receiver operating characteristic curve (ROC-AUC) and the precision-recall curve (PR-AUC), the Brier score, and the calibration slope and intercept (calibration-in-the-large; Supplementary Figure S3). For threshold-based metrics (sensitivity, specificity, and positive and negative predictive values), the threshold maximizing Youden’s *J* (Youden, 1950) was selected from out-of-fold predictions within each training set and then applied to the corresponding test fold. Mean performance and differences between models are reported with 95% confidence intervals from the corrected resampled variance and *t*-test (Nadeau & Bengio, 2003), which account for the overlap between training sets.

#### Hypothesis tests

The nonlinearity hypothesis was tested by comparing the additive EBM with elastic-net logistic regression, and the interaction hypothesis by comparing the EBM with and without interaction terms. Both comparisons test whether nonlinear shapes or interactions improve out-of-sample discrimination, not whether the underlying associations are linear or additive, so the shape functions of the hypothesized features were also examined descriptively.

#### Interpretive model

For interpretation, the EBM was refit to the full sample with the hyperparameters selected most often in cross-validation. Before the analyses were run, we specified that the interaction model would be interpreted only if it outperformed the additive model, that is, if the 95% confidence interval for the difference in PR-AUC excluded zero.

Otherwise, the additive model was interpreted. We report each feature’s shape function, defined as its contribution to the log-odds of SI relative to the sample average, and its importance, defined as its mean absolute contribution. Stability was assessed from 200 refits on stratified 80% subsamples; the resulting bands are heuristic, describing how much a curve varies across refits rather than serving as confidence intervals (Supplementary Section S3). For interaction terms, we report how often each pair was selected and ranked among the three most important.

**Secondary analyses.** Four analyses addressed how to interpret the profile. First, to compare the features with depression severity, models containing BDI-II severity (excluding item 9) alone and together with the 15 features were evaluated in the same nested cross-validation. Depression severity alone was modeled with unpenalized logistic regression and an additive EBM, and the combined models with elastic net and an additive EBM (Supplementary Section S1). To examine whether the two approaches identified the same participants, we compared their out-of-fold predictions (Spearman correlation) and threshold-based classifications (percentage agreement and Cohen’s κ). The analysis was repeated with PROMIS Depression as a second depression measure (Supplementary Table S2).

Second, as a sensitivity analysis, we removed items that might duplicate specific depressive symptoms, using a procedure that did not involve the outcome. For each item of the non-depression scales and each BDI-II item (excluding item 9), we computed the partial correlation controlling for the rest of the item’s own subscale and the BDI-II rest score. These controls absorb general overlap, so a large remaining association suggests, but does not establish, duplicated content. Items with any partial correlation significant after Bonferroni correction across the 4,340 pairs tested were flagged, and flagging was repeated in 500 bootstrap resamples. Items flagged in at least 90% (strict) or 60% (liberal) of resamples were removed, the affected scales were rescored, and the comparisons were repeated (Supplementary Section S2). Because items were selected once in the full sample, before cross-validation, this analysis is exploratory.

Third, because depression-related content can span an entire subscale, conscientiousness was replaced by its two aspects, and the EBM was refit with and without depression severity.

Fourth, to describe associations with conventional inference, we fit Firth-penalized logistic regressions (Firth, 1993) to the full sample for each feature alone, the 15 features together, and the 15 features with depression severity, reporting odds ratios (ORs) per standard deviation with profile penalized-likelihood confidence intervals and tests, in which each coefficient’s restricted fit retained the full model’s penalty (Heinze & Schemper, 2002). Global comparisons of nested models used likelihood-ratio tests of ordinary (unpenalized) maximum-likelihood fits, including a test of whether a general-distress score, the first principal component of the seven standardized non-depression clinical scales, added to depression severity. Because the models used a single imputed dataset, complete-case estimates were examined as a sensitivity analysis. The analyses are exploratory, with *p* values unadjusted for multiple comparisons (Supplementary Section S4).

#### Software and reporting

Analyses used Python 3.11.8 with pandas 2.2.2 (McKinney, 2010), NumPy 1.26.4, SciPy 1.13.1, scikit-learn 1.8.0 (Pedregosa et al., 2011), interpret 0.7.6, and Matplotlib 3.9.2; the Firth models and stability bands used custom code. The study was not preregistered, and no protocol was prepared; its hypotheses and primary analyses were described previously (Zaboski, Mattera, et al., 2026). Reporting follows the TRIPOD+AI statement (Collins et al., 2024), and a completed checklist appears in Supplementary Table S10. The data cannot be shared publicly but are available from the corresponding author on reasonable request. The analysis code is available at https://doi.org/10.5281/zenodo.23028569. Patients and the public were not involved in the design, conduct, reporting, or dissemination of this research.

## Results

### Sample Characteristics

Participants had a mean age of 30.6 years (*SD* = 11.1) and were predominantly female (70.6% by sex; 61.9% by gender identity, with 8.2% identifying as non-binary or another gender), White (77.5%), and not Hispanic or Latino (89.2%). Most (60.6%) reported taking medication. The mean BDI-II total fell in the mild range (*M* = 19.0, *SD* = 11.7), and OCD symptom severity varied widely (DOCS range 2–77). Sixty-six participants (28.6%) endorsed SI. Of these, 61 chose the lowest positive response (thoughts without intent to act), 5 chose the next, and none chose the highest, consistent with the exclusion of active SI with a plan or intent. Table 1 summarizes the sample.

**Table 1.**
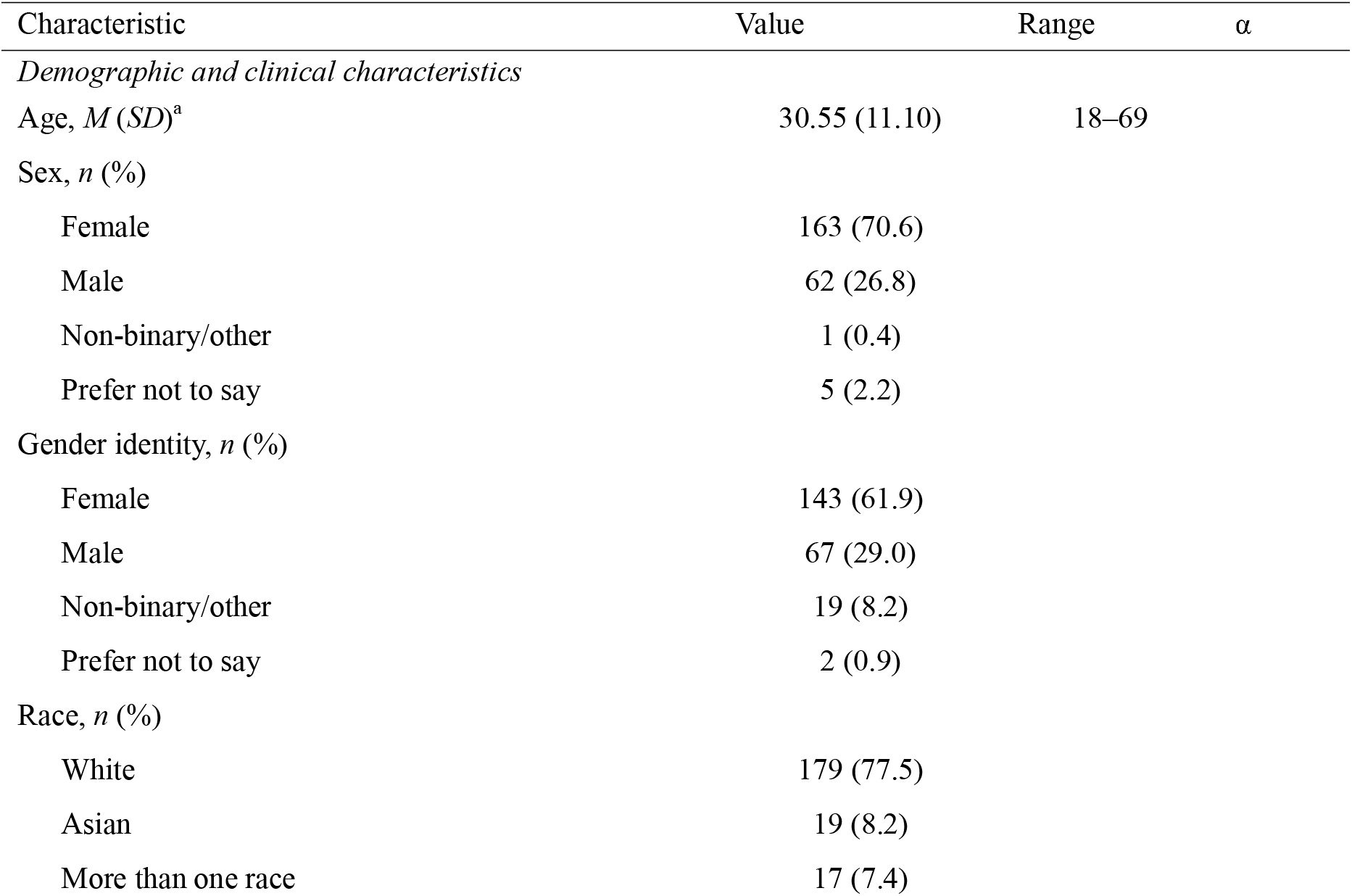
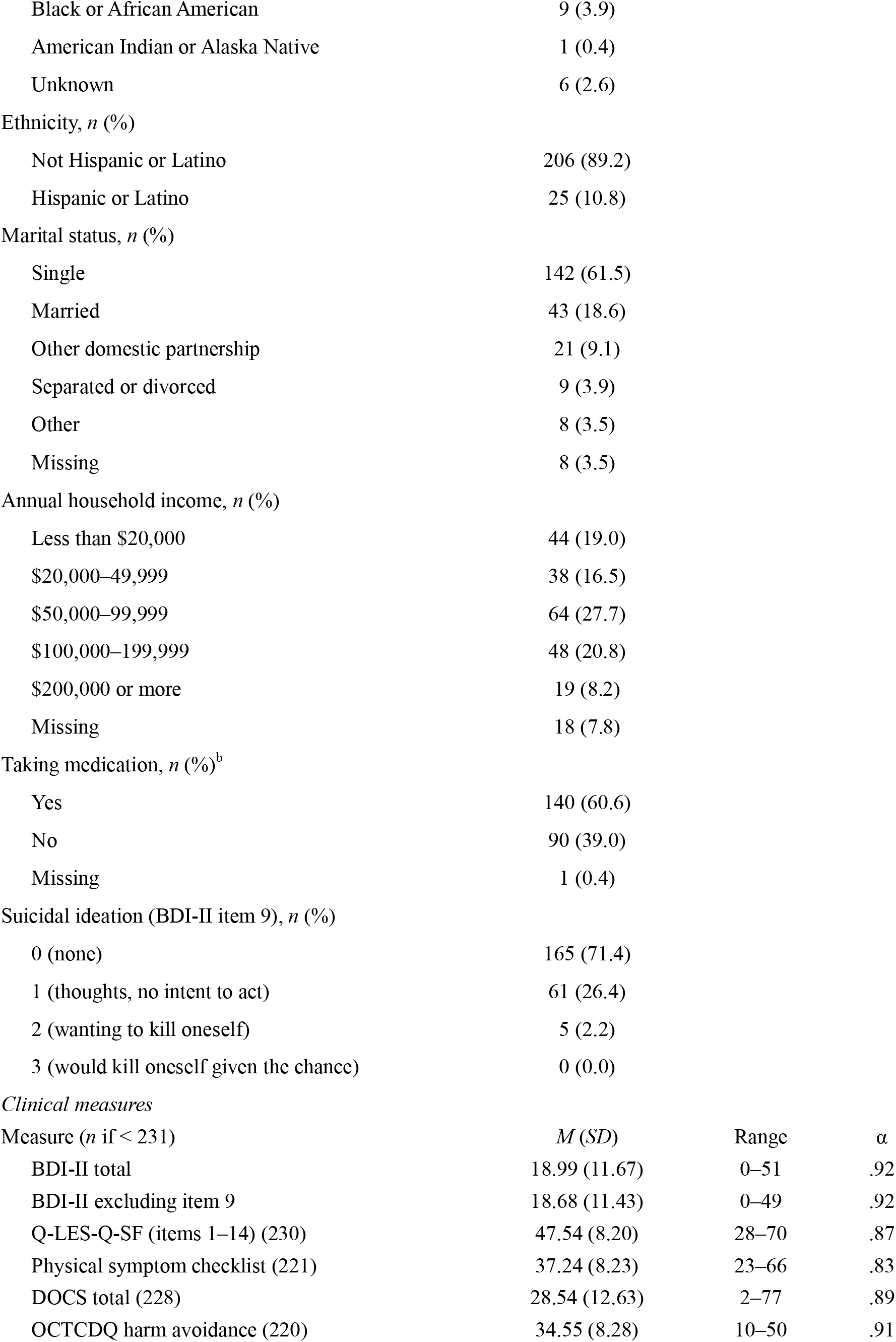
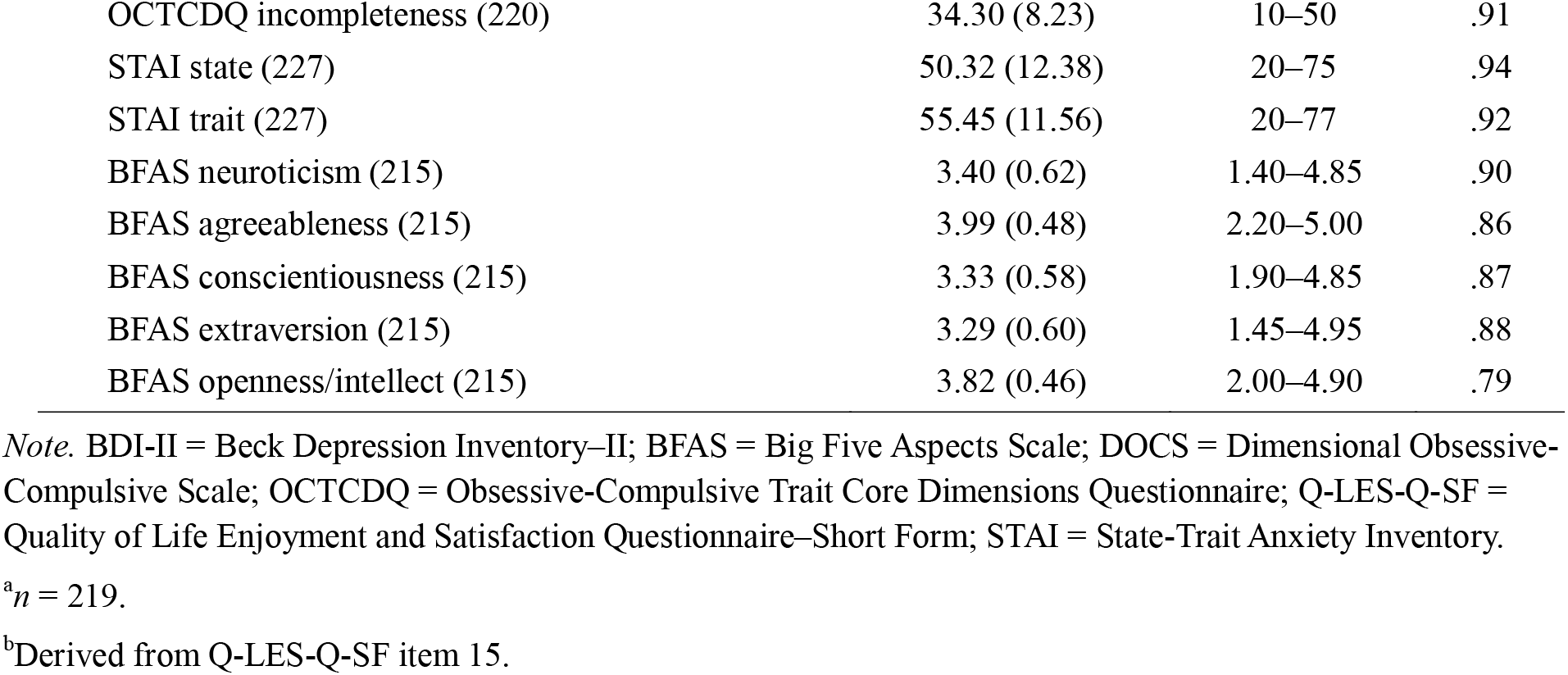
Sample Characteristics (N = 231)

| Characteristic | Value | Range | $\alpha$ |
| --- | --- | --- | --- |
| *Demographic and clinical characteristics* |  |  |  |
| Age, $M$ ($SD$)<sup>a</sup> | 30.55 (11.10) | 18–69 | |
| Sex, $n$ (%) | | | |
| Female | 163 (70.6) |  |  |
| Male | 62 (26.8) |  |  |
| Non-binary/other | 1 (0.4) |  |  |
| Prefer not to say | 5 (2.2) |  |  |
| Gender identity, $n$ (%) | | | |
| Female | 143 (61.9) |  |  |
| Male | 67 (29.0) |  |  |
| Non-binary/other | 19 (8.2) |  |  |
| Prefer not to say | 2 (0.9) |  |  |
| Race, $n$ (%) | | | |
| White | 179 (77.5) |  |  |
| Asian | 19 (8.2) |  |  |
| More than one race | 17 (7.4) |  |  |
| Black or African American | 9 (3.9) |  |  |
| American Indian or Alaska Native | 1 (0.4) |  |  |
| Unknown | 6 (2.6) |  |  |
| Ethnicity, *n* (%) |  |  |  |
| Not Hispanic or Latino | 206 (89.2) |  |  |
| Hispanic or Latino | 25 (10.8) |  |  |
| Marital status, *n* (%) |  |  |  |
| Single | 142 (61.5) |  |  |
| Married | 43 (18.6) |  |  |
| Other domestic partnership | 21 (9.1) |  |  |
| Separated or divorced | 9 (3.9) |  |  |
| Other | 8 (3.5) |  |  |
| Missing | 8 (3.5) |  |  |
| Annual household income, *n* (%) |  |  |  |
| Less than \$20,000 | 44 (19.0) | | |
| \$20,000–49,999 | 38 (16.5) | | |
| \$50,000–99,999 | 64 (27.7) | | |
| \$100,000–199,999 | 48 (20.8) | | |
| \$200,000 or more | 19 (8.2) | | |
| Missing | 18 (7.8) |  |  |
| Taking medication, *n* (%)<sup>b</sup> |  |  |  |
| Yes | 140 (60.6) |  |  |
| No | 90 (39.0) |  |  |
| Missing | 1 (0.4) |  |  |
| Suicidal ideation (BDI-II item 9), *n* (%) |  |  |  |
| 0 (none) | 165 (71.4) |  |  |
| 1 (thoughts, no intent to act) | 61 (26.4) |  |  |
| 2 (wanting to kill oneself) | 5 (2.2) |  |  |
| 3 (would kill oneself given the chance) | 0 (0.0) |  |  |
| *Clinical measures* |  |  |  |
| Measure (*n* if < 231) | *M* (*SD*) | Range | *α* |
| BDI-II total | 18.99 (11.67) | 0–51 | .92 |
| BDI-II excluding item 9 | 18.68 (11.43) | 0–49 | .92 |
| Q-LES-Q-SF (items 1–14) (230) | 47.54 (8.20) | 28–70 | .87 |
| Physical symptom checklist (221) | 37.24 (8.23) | 23–66 | .83 |
| DOCS total (228) | 28.54 (12.63) | 2–77 | .89 |
| OCTCDQ harm avoidance (220) | 34.55 (8.28) | 10–50 | .91 |
| OCTCDQ incompleteness (220) | 34.30 (8.23) | 10–50 | .91 |
| STAI state (227) | 50.32 (12.38) | 20–75 | .94 |
| STAI trait (227) | 55.45 (11.56) | 20–77 | .92 |
| BFAS neuroticism (215) | 3.40 (0.62) | 1.40–4.85 | .90 |
| BFAS agreeableness (215) | 3.99 (0.48) | 2.20–5.00 | .86 |
| BFAS conscientiousness (215) | 3.33 (0.58) | 1.90–4.85 | .87 |
| BFAS extraversion (215) | 3.29 (0.60) | 1.45–4.95 | .88 |
| BFAS openness/intellect (215) | 3.82 (0.46) | 2.00–4.90 | .79 |

*Note.* BDI-II = Beck Depression Inventory–II; BFAS = Big Five Aspects Scale; DOCS = Dimensional Obsessive-Compulsive Scale; OCTCDQ = Obsessive-Compulsive Trait Core Dimensions Questionnaire; Q-LES-Q-SF = Quality of Life Enjoyment and Satisfaction Questionnaire–Short Form; STAI = State-Trait Anxiety Inventory.
<sup>a</sup>*n* = 219.
<sup>b</sup>Derived from Q-LES-Q-SF item 15.

### Model Performance

The four models showed similar discrimination (Table 2), with mean ROC-AUCs of .77– .78 and PR-AUCs of .57–.61 against a base rate of .29. Their confidence intervals were wide, spanning about ±.07 for ROC-AUC and ±.10 to ±.12 for PR-AUC. At thresholds selected within training folds, sensitivity was .65–.73, specificity .70–.77, and positive and negative predictive values .49–.53 and .85–.87. Elastic net, random forest, and the additive EBM were reasonably well calibrated (slopes = 0.96–1.18; intercepts = −0.01 to 0.02), whereas the EBM with interactions was overconfident (slope = 0.62; Supplementary Figure S3).

**Table 2.**
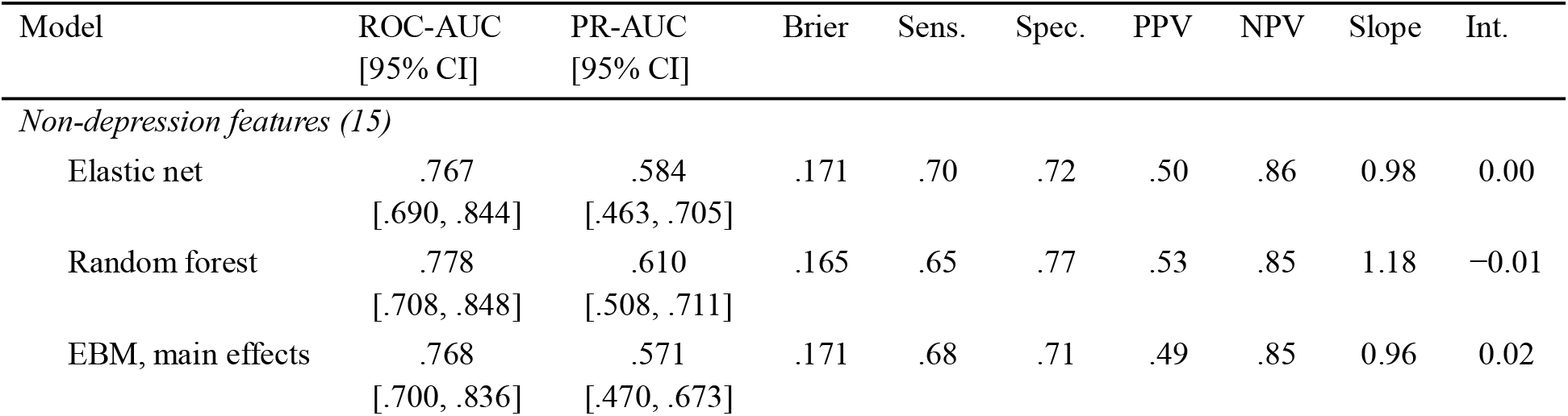
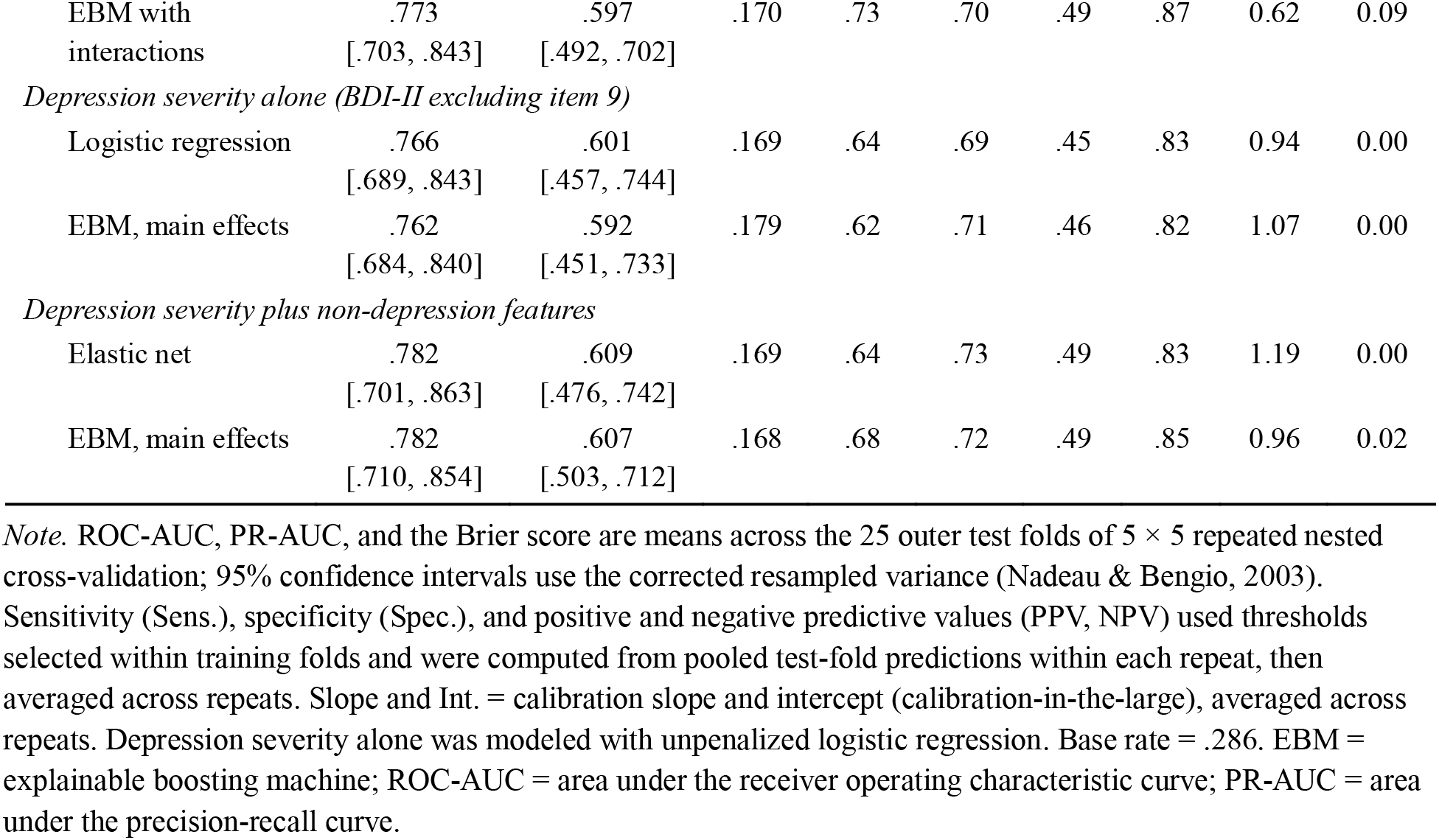
Cross-Validated Performance in Identifying Suicidal Ideation.

| Model | ROC-AUC<br>[95% CI] | PR-AUC<br>[95% CI] | Brier | Sens. | Spec. | PPV | NPV | Slope | Int. |
| --- | --- | --- | --- | --- | --- | --- | --- | --- | --- |
| *Non-depression features (15)* |  |  |  |  |  |  |  |  |  |
| Elastic net | .767<br>[.690, .844] | .584<br>[.463, .705] | .171 | .70 | .72 | .50 | .86 | 0.98 | 0.00 |
| Random forest | .778<br>[.708, .848] | .610<br>[.508, .711] | .165 | .65 | .77 | .53 | .85 | 1.18 | $-0.01$ |
| EBM, main effects | .768<br>[.700, .836] | .571<br>[.470, .673] | .171 | .68 | .71 | .49 | .85 | 0.96 | 0.02 |
| EBM with interactions | .773<br>[.703, .843] | .597<br>[.492, .702] | .170 | .73 | .70 | .49 | .87 | 0.62 | 0.09 |
| *Depression severity alone (BDI-II excluding item 9)* |  |  |  |  |  |  |  |  |  |
| Logistic regression | .766<br>[.689, .843] | .601<br>[.457, .744] | .169 | .64 | .69 | .45 | .83 | 0.94 | 0.00 |
| EBM, main effects | .762<br>[.684, .840] | .592<br>[.451, .733] | .179 | .62 | .71 | .46 | .82 | 1.07 | 0.00 |
| *Depression severity plus non-depression features* |  |  |  |  |  |  |  |  |  |
| Elastic net | .782<br>[.701, .863] | .609<br>[.476, .742] | .169 | .64 | .73 | .49 | .83 | 1.19 | 0.00 |
| EBM, main effects | .782<br>[.710, .854] | .607<br>[.503, .712] | .168 | .68 | .72 | .49 | .85 | 0.96 | 0.02 |

Note. ROC-AUC, PR-AUC, and the Brier score are means across the 25 outer test folds of $5 \times 5$ repeated nested cross-validation; 95% confidence intervals use the corrected resampled variance (Nadeau & Bengio, 2003). Sensitivity (Sens.), specificity (Spec.), and positive and negative predictive values (PPV, NPV) used thresholds selected within training folds and were computed from pooled test-fold predictions within each repeat, then averaged across repeats. Slope and Int. = calibration slope and intercept (calibration-in-the-large), averaged across repeats. Depression severity alone was modeled with unpenalized logistic regression. Base rate = .286. EBM = explainable boosting machine; ROC-AUC = area under the receiver operating characteristic curve; PR-AUC = area under the precision-recall curve.

### Hypothesis Tests

Allowing nonlinear shape functions did not reliably improve discrimination: the additive EBM performed similarly to elastic-net logistic regression (ΔROC-AUC = .001, 95% CI [−.041, .042]; ΔPR-AUC = −.013, 95% CI [−.122, .096]). Adding interaction terms produced no clear gain over the additive EBM (ΔROC-AUC = .005, 95% CI [−.022, .032]; ΔPR-AUC = .026, 95% CI [−.020, .071]), and random forest performed similarly to elastic net (ΔROC-AUC = .011, 95% CI [−.034, .056]). The PR-AUC intervals were too wide to rule out meaningful differences in either direction. Under the rule specified in advance, the additive EBM was interpreted (Table 3).

**Table 3.**
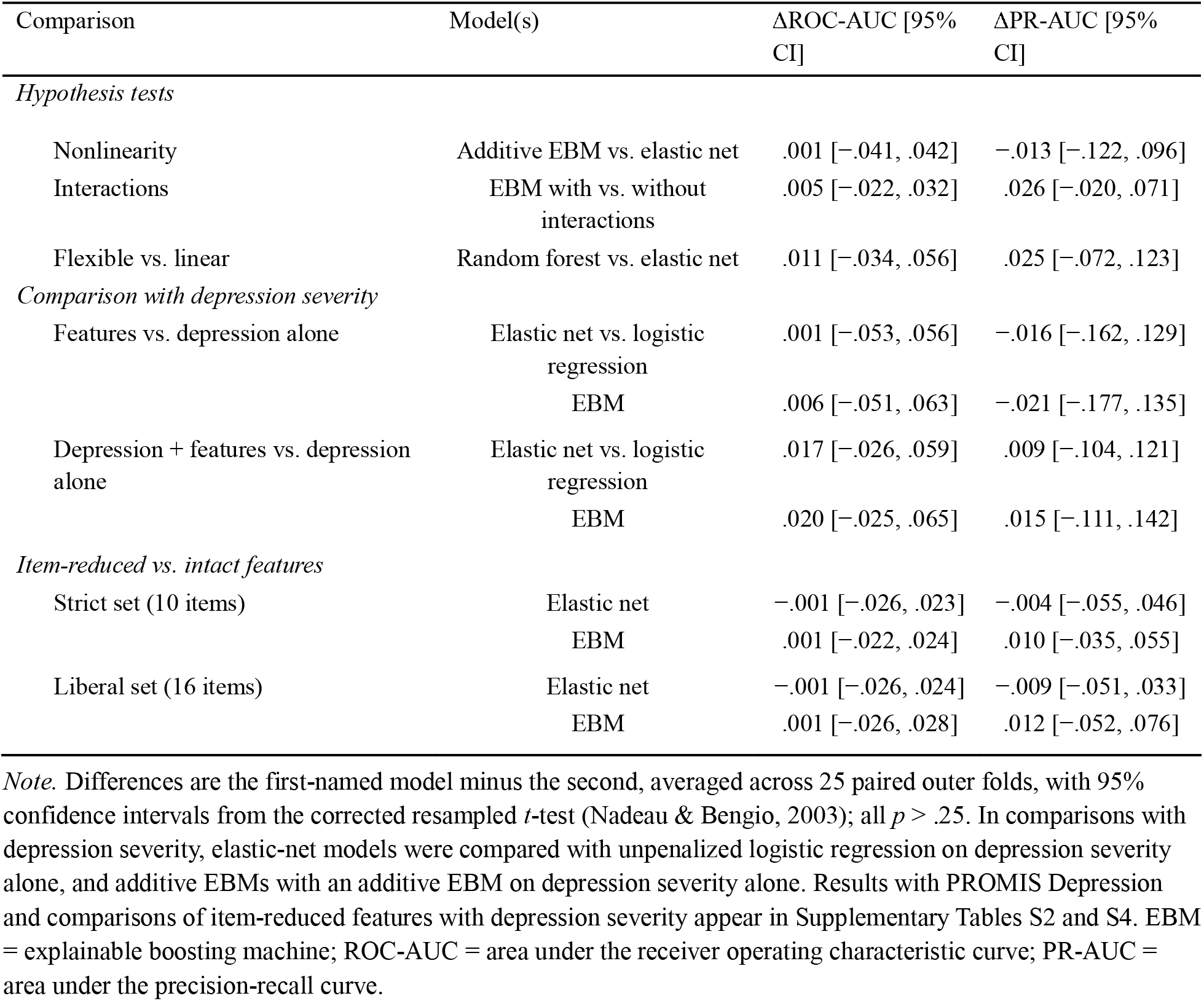
Planned and Secondary Model Comparisons.

| Comparison | Model(s) | ΔROC-AUC [95% CI] | ΔPR-AUC [95% CI] |
| --- | --- | --- | --- |
| *Hypothesis tests* |  |  |  |
| Nonlinearity | Additive EBM vs. elastic net | .001 [−.041, .042] | −.013 [−.122, .096] |
| Interactions | EBM with vs. without interactions | .005 [−.022, .032] | .026 [−.020, .071] |
| Flexible vs. linear | Random forest vs. elastic net | .011 [−.034, .056] | .025 [−.072, .123] |
| *Comparison with depression severity* |  |  |  |
| Features vs. depression alone | Elastic net vs. logistic regression | .001 [−.053, .056] | −.016 [−.162, .129] |
|  | EBM | .006 [−.051, .063] | −.021 [−.177, .135] |
| Depression + features vs. depression alone | Elastic net vs. logistic regression | .017 [−.026, .059] | .009 [−.104, .121] |
|  | EBM | .020 [−.025, .065] | .015 [−.111, .142] |
| *Item-reduced vs. intact features* |  |  |  |
| Strict set (10 items) | Elastic net | −.001 [−.026, .023] | −.004 [−.055, .046] |
|  | EBM | .001 [−.022, .024] | .010 [−.035, .055] |
| Liberal set (16 items) | Elastic net | −.001 [−.026, .024] | −.009 [−.051, .033] |
|  | EBM | .001 [−.026, .028] | .012 [−.052, .076] |

*Note.* Differences are the first-named model minus the second, averaged across 25 paired outer folds, with 95% confidence intervals from the corrected resampled *t*-test (Nadeau & Bengio, 2003); all $p > .25$ . In comparisons with depression severity, elastic-net models were compared with unpenalized logistic regression on depression severity alone, and additive EBMs with an additive EBM on depression severity alone. Results with PROMIS Depression and comparisons of item-reduced features with depression severity appear in Supplementary Tables S2 and S4. EBM = explainable boosting machine; ROC-AUC = area under the receiver operating characteristic curve; PR-AUC = area under the precision-recall curve.

### Comparison With Depression Severity

Depression severity alone and the 15 non-depression features showed similar discrimination (Table 2; ROC-AUC = .766 for depression severity vs. .767 for the features, difference = .001, 95% CI [−.053, .056]; EBM: .762 vs. .768, difference = .006, 95% CI [−.051, .063]). Adding the features to depression severity produced small estimated gains (elastic net: ΔROC-AUC = .017, 95% CI [−.026, .059]; EBM: .020, 95% CI [−.025, .065]). The ROC-AUC intervals are compatible with at most modest gains, up to about .06–.07; the PR-AUC intervals were too wide to bound them (e.g., EBM: ΔPR-AUC = .015, 95% CI [−.111, .142]), largely because each test fold held only 13–14 participants with SI. PROMIS Depression yielded the same pattern (Supplementary Table S2).

In an exploratory in-sample comparison, adding the features improved model fit (likelihood-ratio χ²[15] = 33.7, *p* = .004), but this was not accompanied by a clearly demonstrated improvement in cross-validated discrimination. Similar discrimination did not mean identical predictions. Averaged across the five cross-validation repeats, out-of-fold predictions from depression-only logistic regression and from the elastic net with the 15 non-depression features correlated at ρ = .71, and the two models classified 80% of participants the same way (κ = .59). Among participants with SI, an average of 57% were flagged by both models, 7% by the depression model only, 13% by the feature model only, and 23% by neither model. Out-of-fold predictions from depression severity alone and from the features correlated at ρ = .71, and the two models classified 80% of participants the same way (κ = .59). Of the participants with SI, 57% were flagged by both, 7% by depression severity only, 13% by the features only, and 23% by neither. SI was not confined to participants with elevated depressive symptoms: 9% of those with minimal BDI-II totals (0–13; 8 of 86) endorsed SI, compared with 22%, 32%, and 63% in the mild, moderate, and severe ranges, and these 8 made up 12% of participants with SI.

### Item Overlap With Depression

The outcome-free procedure flagged 16 items whose partial correlation with a specific BDI-II item survived Bonferroni correction in at least 60% of bootstrap resamples, 10 of them in at least 90%, mostly from the STAI trait scale (6 items; e.g., feeling like a failure) and the physical symptom checklist (4; Supplementary Table S3). Not every pairing was an obvious duplication, though. For example, an industriousness item about keeping one’s mind on a task paired most strongly with self-criticalness. Removing the items left discrimination essentially unchanged (ΔROC-AUC from −.001 to .001, all 95% CIs within ±.03; Table 3 and Supplementary Table S4). Because item-reduced and intact scores correlated .97–.99, this shows robustness to removing these items rather than the absence of measurement overlap.

### Interpretive Model

Figure 1 shows the additive EBM’s shape functions. The full-sample fit was optimistic (in-sample ROC-AUC = .90 vs. .77 in cross-validation), so the curves are best read descriptively, and their stability bands were wide. Quality of life made the largest contribution (importance = .34), falling from 0.57 at the 10th percentile to −0.45 at the 90th, and ranked first in all six full-sample specifications examined; the next six features (conscientiousness, trait anxiety, incompleteness, medication use, physical symptoms, and age; importance = .18–.24) appeared in an unstable order (Supplementary Figure S1 and Table S6). Some curves showed steps that the data are too sparse to confirm, notably a drop in the conscientiousness curve near a score of 4.0. The 13% of participants at or above that score accounted for 43% of conscientiousness’s importance. Added to the model, depression severity ranked fifth, or second when conscientiousness was split, and the other features’ importance changed little, reflecting how EBMs share credit among correlated features.

**Figure 1.**
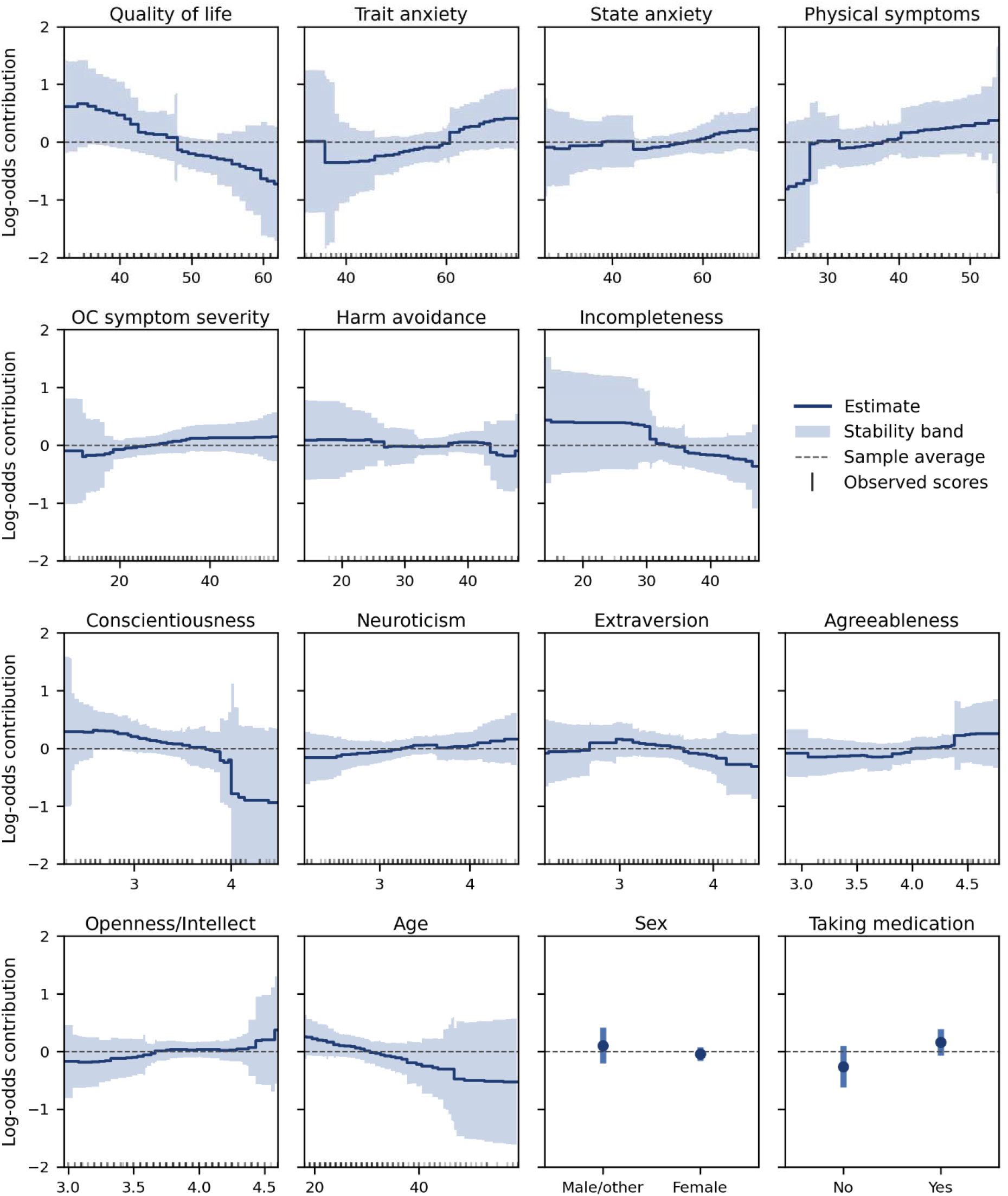
Contributions of each feature to the log-odds of suicidal ideation in the additive explainable boosting machine (0 = sample average), across the central 95% of observed scores. Bands show resampling stability (Supplementary Section S3); ticks mark observed scores.

Splitting conscientiousness divided its contribution between orderliness (importance = .14) and industriousness (.10), both with wide stability bands (Supplementary Figure S2), and adding depression severity reduced neither (Supplementary Table S6). Orderliness was essentially uncorrelated with depression severity (*r* = −.03), whereas industriousness negatively correlated with it (−.48). No interaction term was consistently prominent. The pair most often among the three most important, agreeableness × conscientiousness, reached that rank in 53% of refits of the interaction model; physical symptoms × agreeableness was selected in 84% of refits but ranked in the top three in only 34%; and the hypothesized pairings of physical symptoms with incompleteness and harm avoidance ranked there in 18% and 2% (Supplementary Table S5). The interaction model’s in-sample ROC-AUC of .99 indicated overfitting.

### Conventional Multivariable Models

In conventional models (Supplementary Table S7), most features were associated with SI on their own, including trait anxiety (OR per SD = 2.56, 95% CI [1.79, 3.78]) and OCD symptom severity (1.55 [1.16, 2.08]). Adjusted for one another, these associations attenuated, in several cases substantially, and became less precise (trait anxiety: 1.31 [0.63, 2.89]; OCD symptom severity: 1.35 [0.81, 2.24]). Adjusted for the other non-depression features, lower quality of life (0.47 [0.25, 0.83]) and younger age (0.65 [0.45, 0.91]) were associated with more SI, and higher incompleteness with less (0.60 [0.37, 0.93]), although incompleteness had no association on its own (0.93 [0.70, 1.23]). With depression severity added, the age and incompleteness estimates changed little, whereas the quality-of-life estimate attenuated further and became less certain (0.55 [0.29, 1.01]). Complete-case estimates were similar for these associations, and those for state and trait anxiety shifted within their wide intervals (Supplementary Table S8). A general-distress score, the first principal component of the seven non-depression clinical scales, explained 57% of their variance, correlated with each (|*r*| = .60– .85; Supplementary Table S9), and correlated .78 with depression severity. Adjusted for depression severity, it showed no detectable incremental association, although the interval was wide (OR per SD = 0.99, 95% CI [0.58, 1.67]; likelihood-ratio χ²[1] = 0.00, *p* = .96).

## Discussion

This study asked what clinical features other than depression reveal about SI, meaning endorsement of suicidal thoughts almost always without intent to act, in adults with OCD. Fifteen features spanning functioning, anxiety, physical symptoms, obsessive-compulsive symptoms and traits, and personality identified SI about as well as depression severity did.

Adding them to depression severity improved in-sample fit without a clearly demonstrated gain in cross-validated discrimination, most of their individual associations attenuated under mutual adjustment, and flexible models showed no reliable advantage.

### Distinct Domains, Overlapping Information

The central finding is that clinically distinct domains carried information about SI that overlapped substantially with depression severity. The features and depression severity identified largely but not entirely the same participants, and 13% of participants with SI were identified by the features alone. The attenuation of most associations under mutual adjustment, together with a general-distress component that correlated .78 with depression severity, is consistent with substantial shared variance, as are earlier analyses of overlapping participants from this clinic (Zaboski, Jones, & Mattera, 2025; Zaboski, Jones, Mattera, et al., 2025). The tripartite model suggests that anxiety and depression share a general negative-affect component (Clark & Watson, 1991), and the STAI trait scale itself contains sadness and self-deprecation content alongside anxiety content (Bieling et al., 1998); together, these help explain why trait anxiety was strongly associated with SI on its own, but not once the other features were held constant. Measurement overlap is a second possibility, which the item-reduction analysis could not settle because the reduced scores correlated .97–.99 with the originals. These analyses do not identify a latent construct, so shared distress remains an interpretation rather than an established mechanism.

### Quality of Life

Quality of life was the most consistently prominent feature. It ranked first in every EBM specification, and its association with SI remained evident after adjustment for the other non-depression features, although it attenuated and became less certain once depression severity was added. This extends earlier work in OCD. Among 101 adults entering intensive treatment, those with recent SI reported lower life satisfaction on the same Q-LES-Q-SF, alongside more depressive and OCD symptoms (Storch et al., 2017). Likewise, among 548 outpatients, the SF-36 mental-health domain was among the factors most strongly related to a suicidality continuum, together with depressive severity (Velloso et al., 2016). The present analyses add a joint examination with other clinical domains, a depression benchmark, and a test of whether flexible modeling improves discrimination. Nonetheless, while quality of life was consistently prominent, its association independent of depression was less precisely established. Why dissatisfaction with life accompanies SI is unclear. It could reflect diminished enjoyment (as the tripartite distinction between low positive affect and negative affect would suggest), functional burden, depressive appraisal, or several processes together. It could also accompany the defeat and entrapment described by the IMV model (O’Connor & Kirtley, 2018), but it cannot stand in for measures of them.

### OCD Severity

OCD symptom severity and harm avoidance were associated with SI on their own but contributed little once the other features were held constant, an attenuation seen in several clinical analyses (Angelakis et al., 2015). A small conditional contribution does not mean that OCD burden is unimportant to suicidal processes. If OCD burden contributes to depressive symptoms or to dissatisfaction with life, adjusting for those variables removes information carried along that pathway. Shared vulnerability and measurement overlap are other possibilities, and cross-sectional data cannot distinguish between them. Longitudinal findings make this pathway plausible. In two clinical cohort studies, OCD symptoms predicted later depressive symptoms rather than the reverse (Belli et al., 2023; Tibi et al., 2017), and in a six-year cohort of 325 adults with OCD, greater OCD severity predicted greater SI severity the following year, in models that did not adjust for depression and whose authors proposed depression as a possible mediator (Brown et al., 2019). Thus, our results show what the measured OCD variables added once the other features were known in this sample, not whether OCD matters for SI. They also rest on the DOCS total score, as specific symptom dimensions, which have been linked to suicidality in other samples (Velloso et al., 2016), were not examined separately.

### Nonlinear and Interaction Hypotheses

Comparing model specifications clarified which aspects of the clinical profile were consistent across analyses. Quality of life remained prominent across EBM specifications and was associated with SI after adjustment for the other non-depression features, whereas obsessive-compulsive symptom severity, the focus of the nonlinearity hypothesis, contributed little and had a nearly flat shape function (importance = .09). More specific patterns were less consistent, though. The hypothesized interactions between somatic burden and obsessive-compulsive traits were not reliably prominent across refits, and the apparent conscientiousness threshold occurred in a sparsely represented region of the scale. Allowing nonlinearities and interactions also yielded no clear improvement in discrimination over elastic net. The EBM’s transparency therefore complemented the performance comparisons by showing how individual features contributed to the fitted profile and how those contributions varied across refits.

### Clinical Implications

Information relevant to SI appeared across functioning, anxiety, and physical complaints, not only in depression scores, with 9% of participants with minimal depressive symptoms endorsing SI. Some participants with SI were also flagged by the other features but not by depression severity. Clinicians should therefore ask about SI directly rather than letting depressive symptoms determine whether the question comes up. The models are not screening tools (Franklin et al., 2017); their value is descriptive.

### Limitations and Future Directions

Several limitations qualify these findings. First, SI was measured with a single self-report item and almost always endorsed without intent. Active SI with a plan or intent was exclusionary, so the findings concern low-intent ideation, and item 9 cannot distinguish SI from ego-dystonic intrusive thoughts about suicide (Mattera et al., 2024, 2025). Second, the design was cross-sectional, so directionality and pathways cannot be inferred. Third, the sample came from a single specialty clinic, was predominantly White and female, and was too small to evaluate performance across sociodemographic subgroups. No fairness methods were applied.

Fourth, all measures were self-reported, clinician-rated OCD severity was not available, and several physical symptom items overlap with medication side effects. Fifth, with 66 events, estimates for flexible models and individual features were imprecise; the full-sample EBM was optimistic (in-sample ROC-AUC = .90 vs. .77), so its shape functions are descriptive, and its stability bands are heuristic. Sixth, the item-reduction analysis selected items in the full sample, and the in-sample multivariable models were exploratory, used a single imputation, and had unadjusted *p* values. Prospective studies that measure defeat, entrapment, and suicidal obsessions directly, and that model OCD severity, depression, and quality of life over time, would clarify how these domains relate to the emergence of SI.

## Conclusion

In adults with OCD, clinically distinct domains carried information about SI that overlapped substantially with depression severity, with limited evidence of improved out-of-sample discrimination when these features were added to depression severity. Quality of life was consistently prominent, although its association independent of depression was less precisely established. Associations of total OCD severity and harm avoidance attenuated after adjustment for the other clinical features. Clinicians should ask about SI directly, whatever a patient’s depressive symptoms.

This work received no funding. CP has consulted for Biohaven Pharmaceuticals, Freedom Biosciences, Mind Therapeutics, Loop Bio, and Evecxia Therapeutics; has received research support from Biohaven Pharmaceuticals, Freedom Biosciences, and Transcend

Therapeutics; owns equity in Loop Bio, Mind Therapeutics, and Lucid/Care; receives royalties from Oxford University Press and UpToDate; and holds pending patents on pathogenic antibodies in pediatric OCD, on novel small-molecular therapeutics for OCD, and on novel mechanisms of psychedelic drugs. None of these relationships are relevant to the current work. BAZ and EFM have no conflicts of interest to disclose.

## Data Availability

All data produced in the present study are available upon reasonable request to the authors.

https://doi.org/10.5281/zenodo.23028569

## Supplementary Materials

### Clinical Correlates of Suicidal Ideation in Adults With Obsessive-Compulsive Disorder: An Interpretable Machine Learning Study

#### Contents

S1. Hyperparameter Tuning (Table S1) S2. Item-Overlap Procedure

S3. Resampling Stability Bands S4. Conventional Logistic Models

S5. Supplementary Results (Tables S2–S9; Figures S1–S3) S6. TRIPOD+AI Reporting Checklist (Table S10)

## S1. Hyperparameter Tuning

Hyperparameters were tuned in an inner three-fold cross-validation within each of the 25 outer training folds, using the area under the precision-recall curve as the criterion. Table S1 lists the grids and how often each value was selected. The interpretive models used the most frequently selected combination: 32 bins for the additive EBM, and 10 interaction terms with 32 bins for the interaction model. Depression severity alone was modeled with effectively unpenalized logistic regression (*C* = 10^10^), because a ranking-based tuning criterion cannot select the amount of shrinkage for a single predictor.

**Table S1.** Hyperparameter Grids and Selection Frequency Across 25 Outer Folds.

| Model | Hyperparameter | Values searched | Selected, value (n of 25) |
| --- | --- | --- | --- |
| Elastic-net logistic regression | Inverse regularization strength (C) | 0.01, 0.1, 1, 10 | 0.1 (17), 0.01 (5), 1 (2), 10 (1) |
|  | L1 ratio | 0.1, 0.5, 0.9 | 0.1 (18), 0.5 (5), 0.9 (2) |
|  | Fixed | SAGA solver; 5,000 iterations; no class weighting | — |
| Random forest | Maximum depth | 3, 5, 7 | 7 (11), 5 (10), 3 (4) |
|  | Minimum samples per leaf | 3, 5 | 5 (14), 3 (11) |
|  | Fixed | 300 trees; no class weighting | — |
| EBM, main effects | Maximum bins | 32, 128 | 32 (17), 128 (8) |
|  | Fixed | No interactions; 14 outer bags | — |
| EBM with interactions | Number of interaction terms | 5, 10 | 10 (15), 5 (10) |
|  | Maximum bins | 32, 128 | 32 (18), 128 (7) |
|  | Fixed | 14 outer bags | — |

*Note.* Other settings used the defaults of scikit-learn 1.8.0 and interpret 0.7.6. EBM = explainable boosting machine.

## S2. Item-Overlap Procedure

For each item *j* of a non-depression subscale *S* and each BDI-II item *k* other than item 9, we computed the partial correlation between items *j* and *k*, controlling for the sum of the other items of *S* and the BDI-II total excluding items *k* and 9. The two control scores absorb what an item shares with its own construct and with overall depression severity, so a large remaining association suggests that the two items duplicate specific content, although residual construct overlap or shared method can produce the same result. Subscales were the Q-LES-Q-SF (items 1–14), the physical symptom checklist, the STAI state and trait scales, the 10 BFAS aspects, the OCTCDQ harm avoidance and incompleteness subscales, and the four DOCS dimensions, giving 217 items and 4,340 item pairs. Partial correlations were computed from complete cases for each subscale and tested with Fisher’s *z* (*z* = atanh(*r*)√(*n* − 5)), and an item was flagged when any of its pairs was significant after Bonferroni correction across all 4,340 pairs. The procedure was repeated in 500 bootstrap resamples of participants. Items flagged in at least 60% (liberal) or 90% (strict) of resamples were removed, and the affected scores were recomputed with the same proration rule. These cutoffs follow the range used in stability selection (Meinshausen & Bühlmann, 2010), but that method’s error control assumes half-samples drawn without replacement, so here they are conventions rather than error-controlled thresholds. Selection used the full sample, before cross-validation, and the outcome item was never used. Item-reduced and intact scores correlated .97 to .99, so the analysis tests robustness to removing these items rather than whether measurement overlap has been eliminated.

## S3. Resampling Stability Bands

Shape functions and importance values were refit on 200 stratified subsamples, each containing 80% of participants drawn without replacement. Subsampling was used instead of the bootstrap because duplicated participants would appear on both sides of the EBM’s internal split for early stopping. Each refit’s curves were evaluated on a common grid of 200 points spanning the observed range of each feature and re-centered so that their mean over the full sample’s scores was zero, because each refit is otherwise centered on its own training rows; importance was computed on the same full-sample reference. Because each subsample shares most of its observations with the full sample, the spread of the refits understates sampling variability. For a statistic with sampling variance σ²/*n*, an estimate from a subsample of size *m* drawn without replacement from the observed sample has variance σ²(1/*m* − 1/*n*), which at *m* = 0.8*n* is one quarter of σ²/*n*. Bands were therefore formed as the full-sample estimate ± 1.96 × √(*m*/[*n* − *m*]) × the root-mean-square deviation of the refits from that estimate, where √(*m*/[*n* − *m*]) = 2.

Importance bands were truncated at zero.

These bands are heuristic stability bands, not confidence intervals. A simulation with *n* = 231 checked only the scaling: for a sample mean, the standard deviation across 80% subsamples was 0.49 times the true standard error (theoretical value, 0.50), and for a logistic regression coefficient, averaged over 40 simulated datasets, the ratio was 0.59. These checks concern standard-error ratios for simple statistics; they do not establish coverage for EBM shape functions, which involve learned bins, regularization, early stopping, and correlated features. The refits were also skewed, tending to be more extreme at the tails than the full-sample fit, so percentile-based bands depend on how that skew is handled. Adding the rescaled percentiles of the deviations to the estimate, and reflecting them around it as in subsampling theory (Politis & Romano, 1994), disagreed about whether zero was excluded at the percentiles cited in the main text; at the 90th percentile of trait anxiety, for example, they gave [0.04, 1.01] and [−0.29, 0.69]. The symmetric band avoids that dependence, and systematic displacement widens it rather than shifting it. Because refits share most of their data, refit-based frequencies, such as how often a term ranked among the three most important, also overstate stability; they are used only to show instability.

## S4. Conventional Logistic Models

The Firth-penalized logistic regressions (Firth, 1993) maximized the log-likelihood plus half the log-determinant of the Fisher information, which reduces small-sample bias and keeps estimates finite. They were fit by Newton–Raphson iteration on the modified score, with step-halving. For each coefficient, the profile penalized-likelihood confidence interval and test were obtained by refitting with that coefficient fixed while retaining the full model’s penalty, as in the logistf package (Heinze & Schemper, 2002). Subtracting penalized log-likelihoods from separately fitted models would not give a valid test, because their penalties differ; for quality of life, that naive comparison would have given *p* = .002 instead of .009. The implementation reproduced the closed-form Firth estimate for a 2 × 2 table (the log odds ratio after adding 0.5 to each cell), converged to maximum likelihood in large samples, and matched an independent general-purpose optimizer (e.g., quality of life: OR = 0.47, 95% CI [0.25, 0.83], *p* = .009).

Refitting every model with the logistf package reproduced all odds ratios, profile intervals, and p values to within 10^−6^ (largest discrepancy, 6.4 × 10^−7^ in an odds ratio), and glm reproduced both likelihood-ratio tests; the R script accompanies the code.

Global likelihood-ratio tests compared nested ordinary (unpenalized) maximum-likelihood models fit to the same data, so the difference in log-likelihoods is a standard test statistic. All in-sample models used one dataset in which missing values were imputed by *k*-nearest neighbors (*k* = 5, standardized distances) on the 15 features and depression severity together; continuous variables were then standardized, so ORs are per standard deviation.

Because a single imputation does not propagate uncertainty about missing values, the multivariable models were refit on the 199 complete cases (55 with suicidal ideation), restandardized within that sample (Table S8). The general-distress score was the first principal component of the seven non-depression clinical scales (quality of life, physical symptoms, OC symptom severity, harm avoidance, incompleteness, and state and trait anxiety) after standardization, oriented so that higher scores indicate more distress (Table S9). Agreement between depression severity alone and the 15 features was computed from out-of-fold predictions and threshold-based classifications within each repeat of cross-validation and averaged across repeats.

## S5. Supplementary Results

**Table S2.** Results With PROMIS Depression as the Depression Measure.

| Model | ROC-AUC<br>[95% CI] | PR-AUC<br>[95% CI] | Brier | ΔROC-AUC [95% CI] | ΔPR-AUC [95% CI] |
| --- | --- | --- | --- | --- | --- |
| *PROMIS Depression alone* |  |  |  |  |  |
| Logistic regression | .761<br>[.685, .836] | .583<br>[.471, .694] | .170 | — | — |
| EBM, main effects | .754<br>[.669, .839] | .595<br>[.476, .714] | .174 | — | — |
| *PROMIS Depression plus non-depression features (vs. PROMIS alone)* |  |  |  |  |  |
| Elastic net | .780<br>[.706, .855] | .590<br>[.476, .704] | .169 | .020 [−.027, .066] | .007 [−.118, .132] |
| EBM, main effects | .781<br>[.710, .851] | .597<br>[.497, .697] | .167 | .027 [−.033, .087] | .002 [−.120, .125] |
| *Non-depression features alone (vs. PROMIS alone)* |  |  |  |  |  |
| Elastic net | .767<br>[.690, .844] | .584<br>[.463, .705] | .171 | .007 [−.051, .064] | .001 [−.133, .136] |
| EBM, main effects | .768<br>[.700, .836] | .571<br>[.470, .673] | .171 | .014 [−.055, .083] | −.023 [−.164, .118] |

*Note.* ROC-AUC and PR-AUC are means across 25 outer folds, with 95% confidence intervals from the corrected resampled variance; Brier scores are means. Differences are the listed model minus PROMIS Depression alone (unpenalized logistic regression for elastic-net rows, an additive EBM for EBM rows), with 95% confidence intervals from the corrected resampled *t*-test; all $p > .35$ . PROMIS scores were available for 214 participants; missing values were imputed within training folds. The PROMIS Depression bank includes an item about having no reason for living.

**Table S3.**
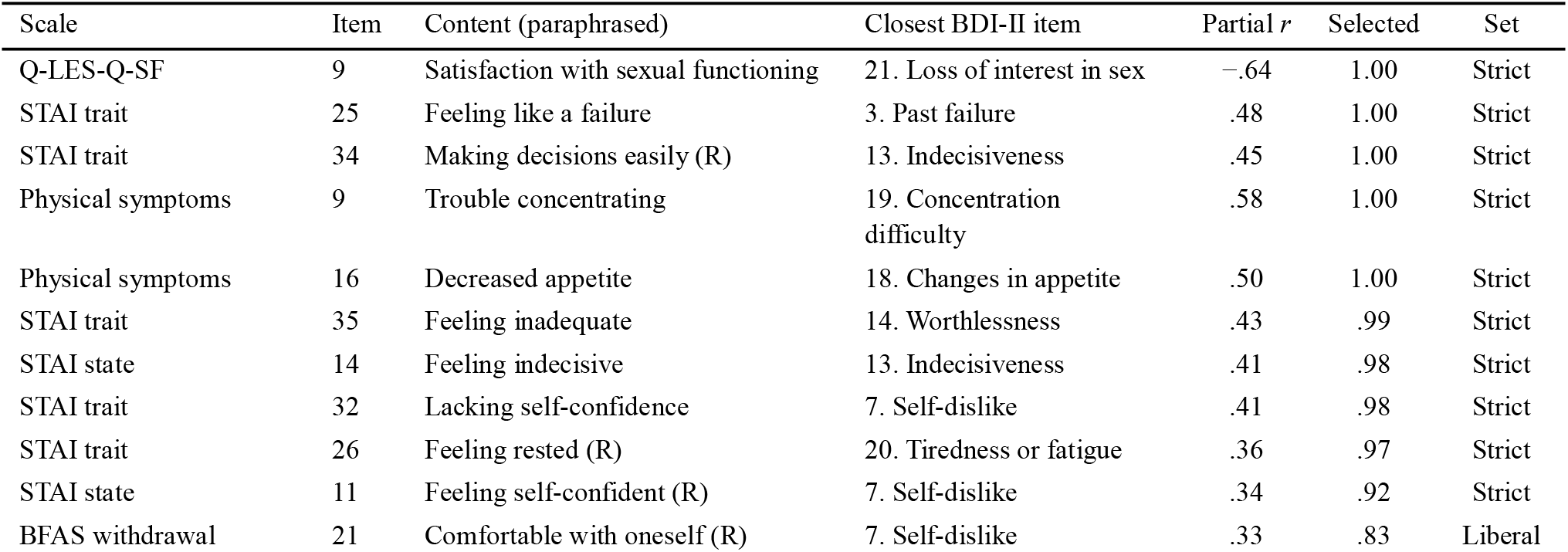
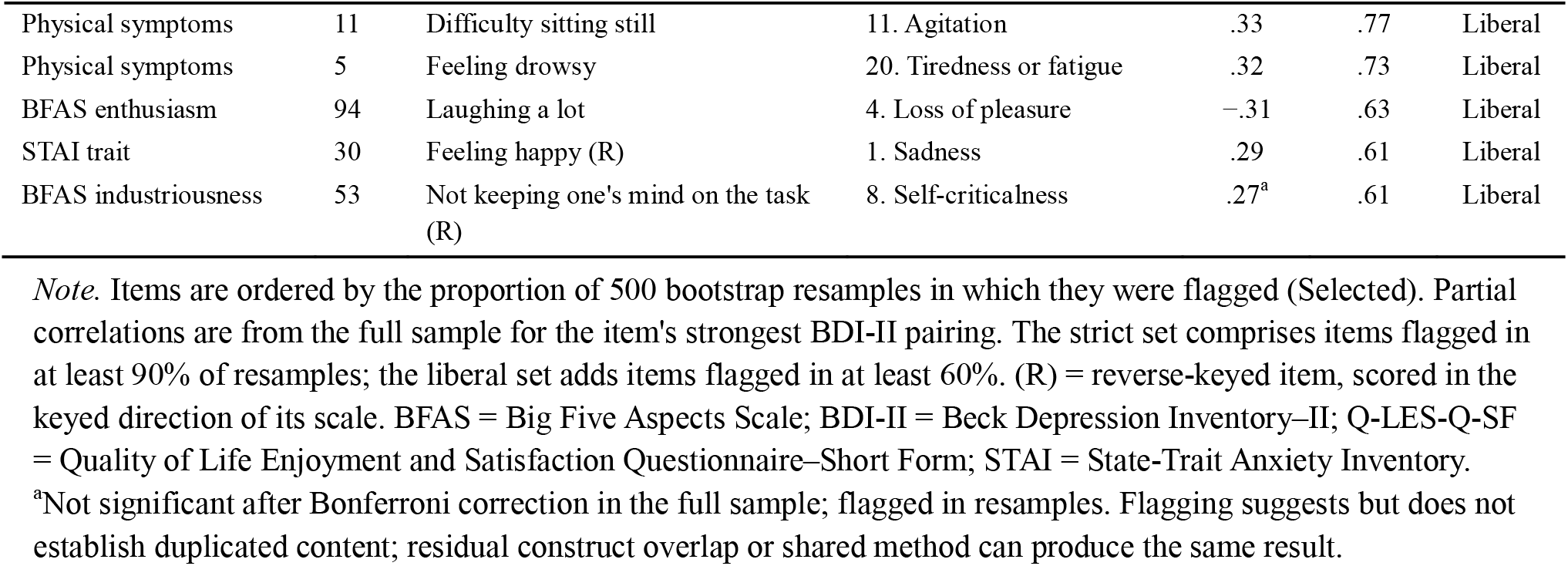
Items Flagged as Possibly Duplicating Specific BDI-II Content.

**Table S4.** Performance of Item-Reduced Feature Sets and Comparisons With Depression Severity.

| Model | ROC-AUC<br>[95% CI] | PR-AUC<br>[95% CI] | Vs. depression<br>alone: ΔROC-AUC | ΔPR-AUC | Added to<br>depression:<br>ΔROC-AUC | ΔPR-AUC |
| --- | --- | --- | --- | --- | --- | --- |
| *Strict set (10 items removed)* |  |  |  |  |  |  |
| Elastic net | .766<br>[.692, .839] | .580<br>[.463, .696] | .000 [–.056,<br>.056] | –.021 [–.169,<br>.127] | .017 [–.025,<br>.058] | –.003 [–.124,<br>.119] |
| EBM, main<br>effects | .769<br>[.700, .838] | .581<br>[.479, .684] | .007 [–.044,<br>.058] | –.011 [–.162,<br>.141] | .016 [–.021,<br>.054] | .011 [–.117,<br>.139] |
| *Liberal set (16 items removed)* |  |  |  |  |  |  |
| Elastic net | .766<br>[.700, .832] | .576<br>[.470, .682] | .001 [–.054,<br>.055] | –.025 [–.169,<br>.119] | .016 [–.025,<br>.057] | .001 [–.120,<br>.122] |
| EBM, main<br>effects | .769<br>[.696, .842] | .583<br>[.484, .682] | .007 [–.043,<br>.058] | –.009 [–.157,<br>.139] | .016 [–.020,<br>.051] | .006 [–.115,<br>.126] |

*Note.* ROC-AUC and PR-AUC are means across 25 outer folds with 95% confidence intervals from the corrected resampled variance. "Vs. depression alone" compares item-reduced features alone with depression severity alone (BDI-II excluding item 9); "added to depression" compares depression severity plus item-reduced features with depression severity alone. Depression severity alone was modeled with unpenalized logistic regression for elastic-net rows and an additive EBM for EBM rows. Differences have 95% confidence intervals from the corrected resampled *t*-test; all *p* > .35. Removing the flagged items changed the scales' correlations with BDI-II severity only slightly (strict/liberal): trait anxiety, .76 to .72/.71; state anxiety, .67 to .66/.66; quality of life, –.75 to –.72/–.72; physical symptoms, .60 to .56/.52; neuroticism, .53 to .51 (liberal only). Item-reduced and intact scores correlated .97 to .99.

**Table S5.**
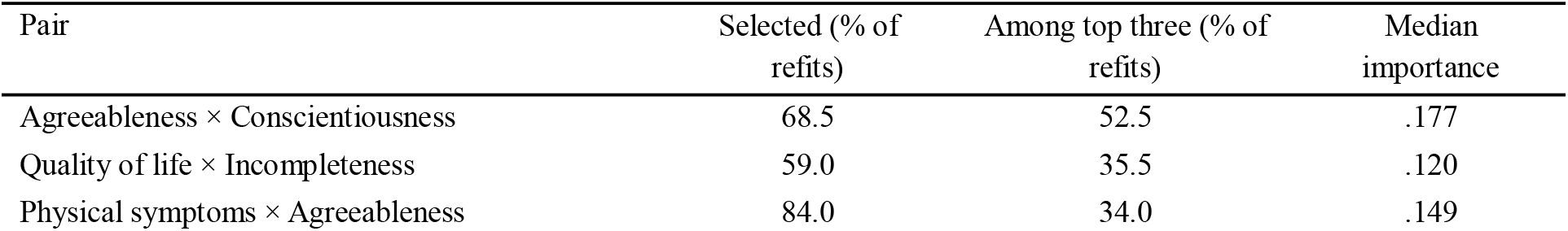
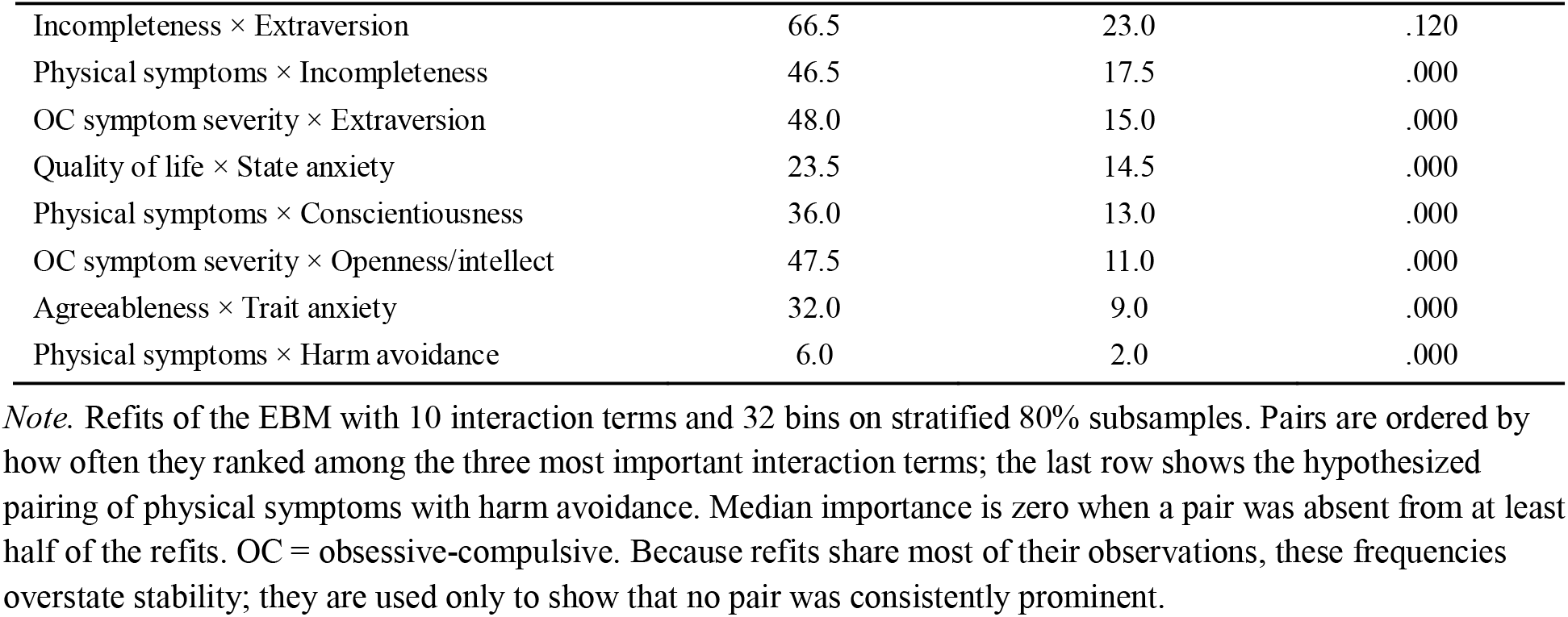
Stability of Interaction Terms Across 200 Subsample Refits.

*Stability of Interaction Terms Across 200 Subsample Refits*
| Pair | Selected (% of<br>refits) | Among top three (% of<br>refits) | Median<br>importance |
| --- | --- | --- | --- |
| Agreeableness × Conscientiousness | 68.5 | 52.5 | .177 |
| Quality of life × Incompleteness | 59.0 | 35.5 | .120 |
| Physical symptoms × Agreeableness | 84.0 | 34.0 | .149 |

OCD, DEPRESSION, AND SUICIDAL IDEATION
| Pair | Selected (% of refits) | Among top three (% of refits) | Median importance |
| --- | --- | --- | --- |
| Incompleteness × Extraversion | 66.5 | 23.0 | .120 |
| Physical symptoms × Incompleteness | 46.5 | 17.5 | .000 |
| OC symptom severity × Extraversion | 48.0 | 15.0 | .000 |
| Quality of life × State anxiety | 23.5 | 14.5 | .000 |
| Physical symptoms × Conscientiousness | 36.0 | 13.0 | .000 |
| OC symptom severity × Openness/intellect | 47.5 | 11.0 | .000 |
| Agreeableness × Trait anxiety | 32.0 | 9.0 | .000 |
| Physical symptoms × Harm avoidance | 6.0 | 2.0 | .000 |

*Note.* Refits of the EBM with 10 interaction terms and 32 bins on stratified 80% subsamples. Pairs are ordered by how often they ranked among the three most important interaction terms; the last row shows the hypothesized pairing of physical symptoms with harm avoidance. Median importance is zero when a pair was absent from at least half of the refits. OC = obsessive-compulsive. Because refits share most of their observations, these frequencies overstate stability; they are used only to show that no pair was consistently prominent.

**Table S6.**
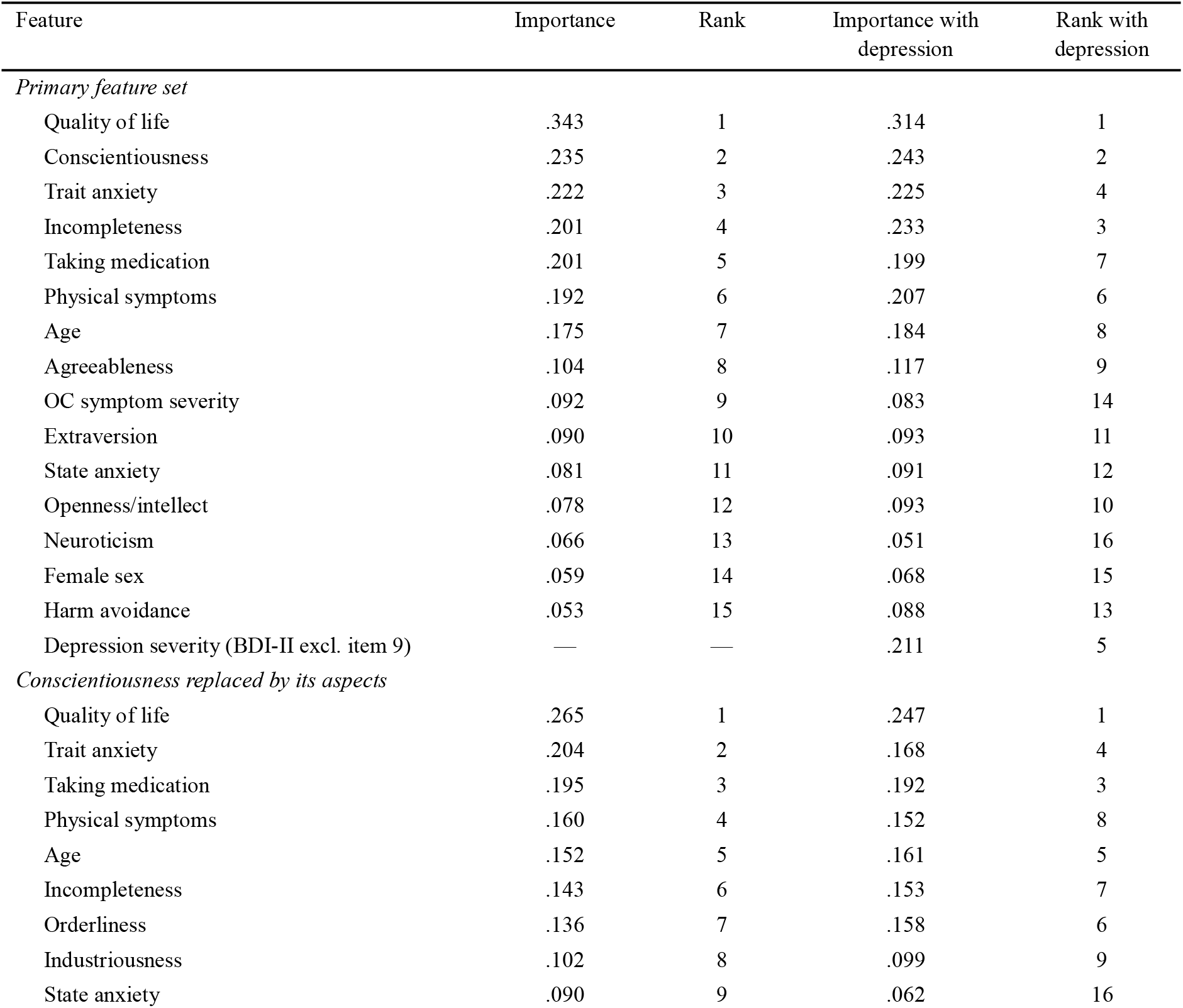
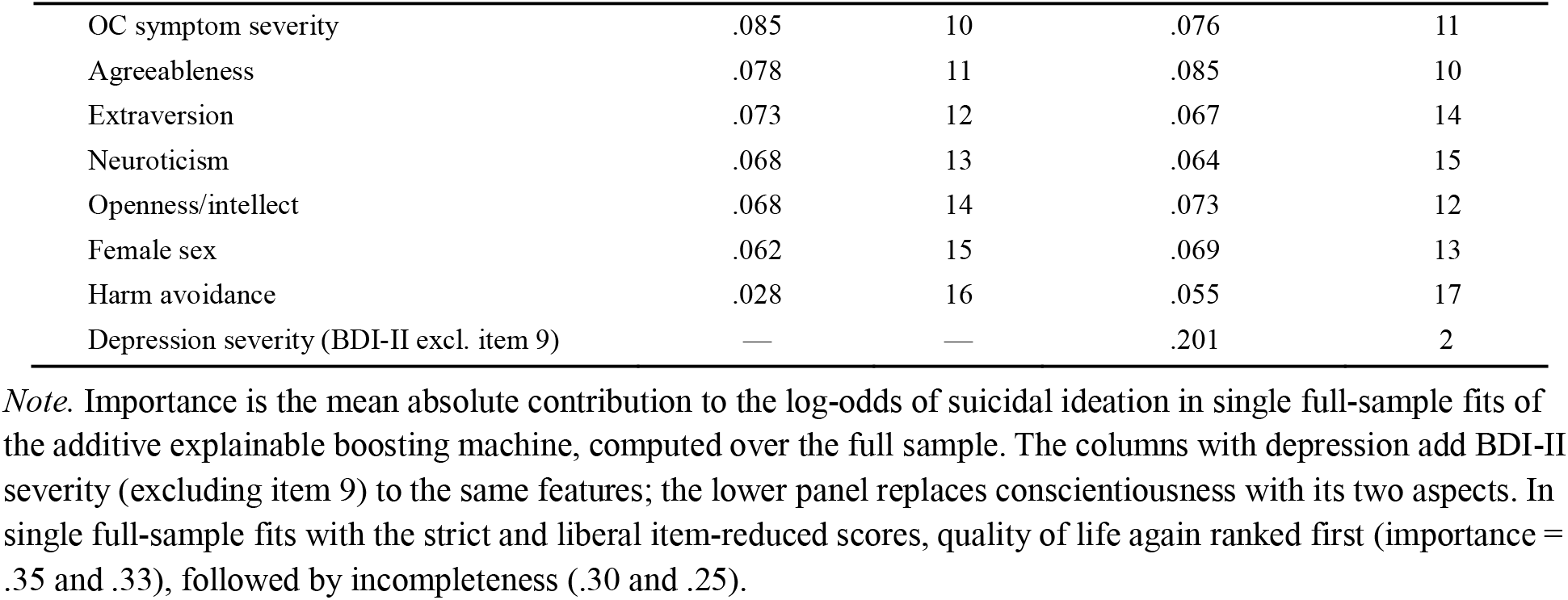
Term Importance With and Without Depression Severity.

*Term Importance With and Without Depression Severity*
| Feature | Importance | Rank | Importance with depression | Rank with depression |
| --- | --- | --- | --- | --- |
| *Primary feature set* |  |  |  |  |
| Quality of life | .343 | 1 | .314 | 1 |
| Conscientiousness | .235 | 2 | .243 | 2 |
| Trait anxiety | .222 | 3 | .225 | 4 |
| Incompleteness | .201 | 4 | .233 | 3 |
| Taking medication | .201 | 5 | .199 | 7 |
| Physical symptoms | .192 | 6 | .207 | 6 |
| Age | .175 | 7 | .184 | 8 |
| Agreeableness | .104 | 8 | .117 | 9 |
| OC symptom severity | .092 | 9 | .083 | 14 |
| Extraversion | .090 | 10 | .093 | 11 |
| State anxiety | .081 | 11 | .091 | 12 |
| Openness/intellect | .078 | 12 | .093 | 10 |
| Neuroticism | .066 | 13 | .051 | 16 |
| Female sex | .059 | 14 | .068 | 15 |
| Harm avoidance | .053 | 15 | .088 | 13 |
| Depression severity (BDI-II excl. item 9) | — | — | .211 | 5 |
| *Conscientiousness replaced by its aspects* |  |  |  |  |
| Quality of life | .265 | 1 | .247 | 1 |
| Trait anxiety | .204 | 2 | .168 | 4 |
| Taking medication | .195 | 3 | .192 | 3 |
| Physical symptoms | .160 | 4 | .152 | 8 |
| Age | .152 | 5 | .161 | 5 |
| Incompleteness | .143 | 6 | .153 | 7 |
| Orderliness | .136 | 7 | .158 | 6 |
| Industriousness | .102 | 8 | .099 | 9 |
| State anxiety | .090 | 9 | .062 | 16 |

OCD, DEPRESSION, AND SUICIDAL IDEATION
| Feature | Importance | Rank | Importance with depression | Rank with depression |
| --- | --- | --- | --- | --- |
| OC symptom severity | .085 | 10 | .076 | 11 |
| Agreeableness | .078 | 11 | .085 | 10 |
| Extraversion | .073 | 12 | .067 | 14 |
| Neuroticism | .068 | 13 | .064 | 15 |
| Openness/intellect | .068 | 14 | .073 | 12 |
| Female sex | .062 | 15 | .069 | 13 |
| Harm avoidance | .028 | 16 | .055 | 17 |
| Depression severity (BDI-II excl. item 9) | — | — | .201 | 2 |

*Note.* Importance is the mean absolute contribution to the log-odds of suicidal ideation in single full-sample fits of the additive explainable boosting machine, computed over the full sample. The columns with depression add BDI-II severity (excluding item 9) to the same features; the lower panel replaces conscientiousness with its two aspects. In single full-sample fits with the strict and liberal item-reduced scores, quality of life again ranked first (importance = .35 and .33), followed by incompleteness (.30 and .25).

**Table S7.**
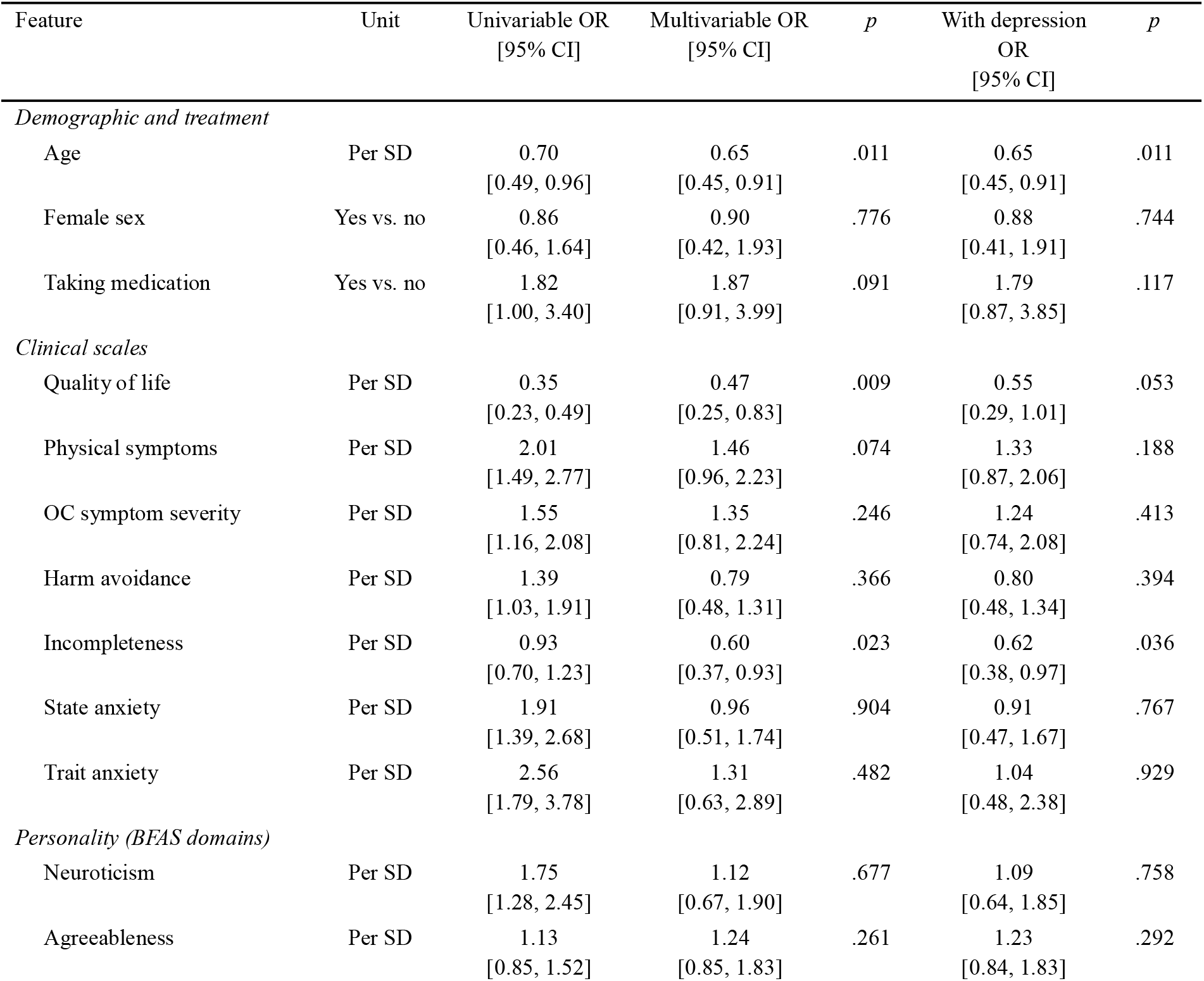
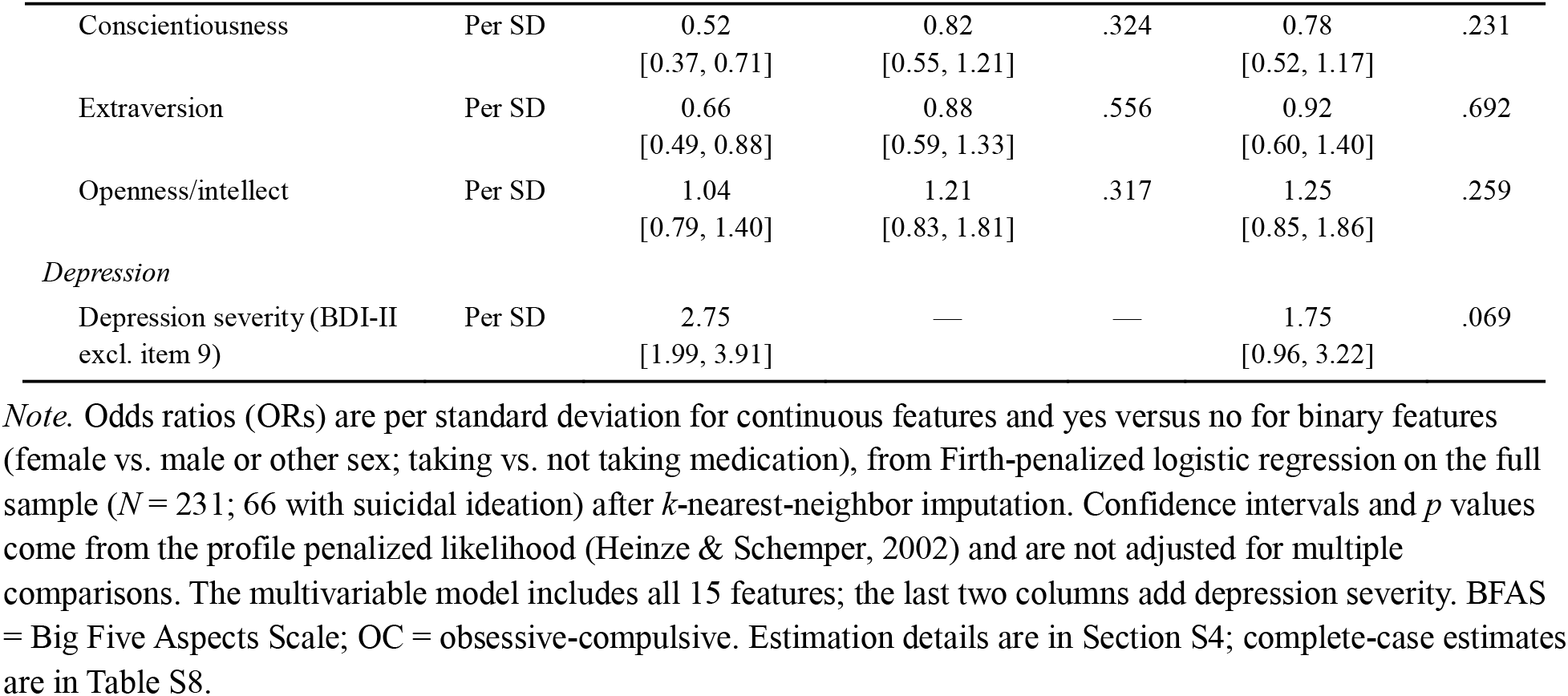
Firth-Penalized Logistic Regressions of Suicidal Ideation.

**Table S8.** Complete-Case Sensitivity Analysis of the Firth-Penalized Multivariable Models.

| Feature | Multivariable OR,<br>imputed<br>[95% CI] | Multivariable OR,<br>complete cases<br>[95% CI] | With depression OR,<br>imputed<br>[95% CI] | With depression OR,<br>complete cases<br>[95% CI] |
| --- | --- | --- | --- | --- |
| *Demographic and treatment* |  |  |  |  |
| Age | 0.65<br>[0.45, 0.91] | 0.70<br>[0.48, 1.00] | 0.65<br>[0.45, 0.91] | 0.71<br>[0.48, 1.00] |
| Female sex | 0.90<br>[0.42, 1.93] | 0.76<br>[0.35, 1.68] | 0.88<br>[0.41, 1.91] | 0.73<br>[0.33, 1.63] |
| Taking medication | 1.87<br>[0.91, 3.99] | 1.88<br>[0.88, 4.19] | 1.79<br>[0.87, 3.85] | 1.82<br>[0.84, 4.10] |
| *Clinical scales* |  |  |  |  |
| Quality of life | 0.47<br>[0.25, 0.83] | 0.46<br>[0.24, 0.84] | 0.55<br>[0.29, 1.01] | 0.55<br>[0.28, 1.02] |
| Physical symptoms | 1.46<br>[0.96, 2.23] | 1.39<br>[0.90, 2.17] | 1.33<br>[0.87, 2.06] | 1.27<br>[0.80, 2.00] |
| OC symptom severity | 1.35<br>[0.81, 2.24] | 1.44<br>[0.84, 2.46] | 1.24<br>[0.74, 2.08] | 1.29<br>[0.75, 2.24] |
| Harm avoidance | 0.79<br>[0.48, 1.31] | 0.70<br>[0.41, 1.17] | 0.80<br>[0.48, 1.34] | 0.70<br>[0.41, 1.20] |
| Incompleteness | 0.60<br>[0.37, 0.93] | 0.59<br>[0.37, 0.93] | 0.62<br>[0.38, 0.97] | 0.60<br>[0.37, 0.95] |
| State anxiety | 0.96<br>[0.51, 1.74] | 0.68<br>[0.34, 1.31] | 0.91<br>[0.47, 1.67] | 0.63<br>[0.31, 1.24] |
| Trait anxiety | 1.31<br>[0.63, 2.89] | 1.54<br>[0.66, 3.77] | 1.04<br>[0.48, 2.38] | 1.24<br>[0.52, 3.09] |
| *Personality (BFAS domains)* |  |  |  |  |
| Neuroticism | 1.12<br>[0.67, 1.90] | 1.29<br>[0.73, 2.32] | 1.09<br>[0.64, 1.85] | 1.25<br>[0.70, 2.27] |

OCD, DEPRESSION, AND SUICIDAL IDEATION
| Feature | Multivariable OR,<br>imputed<br>[95% CI] | Multivariable OR,<br>complete cases<br>[95% CI] | With depression OR,<br>imputed<br>[95% CI] | With depression OR,<br>complete cases<br>[95% CI] |
| --- | --- | --- | --- | --- |
| Agreeableness | 1.24<br>[0.85, 1.83] | 1.21<br>[0.80, 1.86] | 1.23<br>[0.84, 1.83] | 1.19<br>[0.78, 1.84] |
| Conscientiousness | 0.82<br>[0.55, 1.21] | 0.87<br>[0.57, 1.32] | 0.78<br>[0.52, 1.17] | 0.83<br>[0.54, 1.28] |
| Extraversion | 0.88<br>[0.59, 1.33] | 0.87<br>[0.57, 1.33] | 0.92<br>[0.60, 1.40] | 0.90<br>[0.58, 1.39] |
| Openness/intellect | 1.21<br>[0.83, 1.81] | 1.30<br>[0.86, 2.02] | 1.25<br>[0.85, 1.86] | 1.35<br>[0.89, 2.10] |
| *Depression* |  |  |  |  |
| Depression severity (BDI-II<br>excl. item 9) | — | — | 1.75<br>[0.96, 3.22] | 1.84<br>[0.99, 3.47] |

*Note.* Odds ratios (ORs) are per standard deviation for continuous features and yes versus no for binary features, from Firth-penalized logistic regression with profile penalized-likelihood 95% confidence intervals. Imputed estimates are from Table S7 ( $N = 231$ ; 66 with suicidal ideation). Complete-case estimates use the 199 participants without missing values (55 with suicidal ideation), with continuous features standardized within that sample. BFAS = Big Five Aspects Scale; OC = obsessive-compulsive.

**Table S9.** Composition of the General-Distress Component.

| Scale | Weight | Correlation with component |
| --- | --- | --- |
| Quality of life (Q-LES-Q-SF) | -.38 | -.76 |
| Physical symptoms | .37 | .73 |
| OC symptom severity (DOCS) | .37 | .74 |
| Harm avoidance (OCTCDQ) | .37 | .75 |
| Incompleteness (OCTCDQ) | .30 | .60 |
| State anxiety (STAI) | .42 | .85 |
| Trait anxiety (STAI) | .41 | .83 |

*Note.* The component is the first principal component of the seven scales after $k$ -nearest-neighbor imputation and standardization. Weights are the elements of the unit-length eigenvector, and correlations are between each scale and the component score. The component is oriented so that higher scores indicate more distress; quality of life, on which higher scores indicate better quality of life, therefore loads negatively. The component explained 57% of the scales' variance and correlated .78 with depression severity (BDI-II excluding item 9) and .32 with suicidal ideation. Per standard deviation, its OR for suicidal ideation was 2.30 (95% CI [1.62, 3.27]) alone and 0.99 (95% CI [0.58, 1.67]) adjusted for depression severity (likelihood-ratio $\chi^2[1] = 0.00$ , $p = .96$ ); the corresponding Firth estimate was 0.98 [0.59, 1.66]. DOCS = Dimensional Obsessive-Compulsive Scale; OCTCDQ = Obsessive-Compulsive Trait Core Dimensions Questionnaire; Q-LES-Q-SF = Quality of Life Enjoyment and Satisfaction Questionnaire–Short Form; STAI = State-Trait Anxiety Inventory.

**Figure S1.**
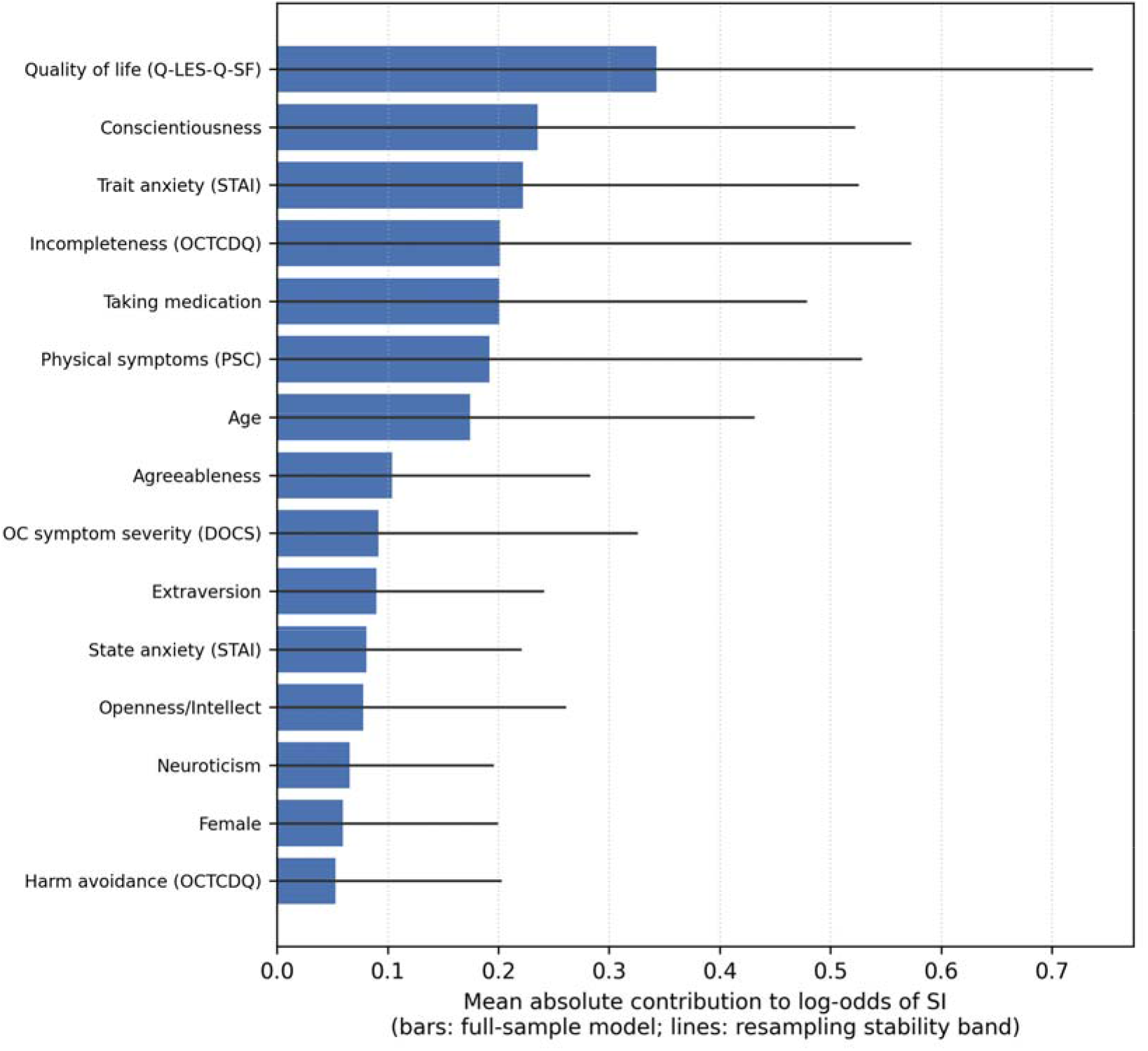
Term Importance in the Additive Explainable Boosting Machine. *Note.* Bars show each feature’s mean absolute contribution to the log-odds of suicidal ideation in the full-sample model; lines show resampling stability bands from rescaled subsample refits, truncated at zero (Section S3). Across refits, only quality of life had a median rank of 1; the other features’ median ranks ranged from 3 to 14.

**Figure S2.**
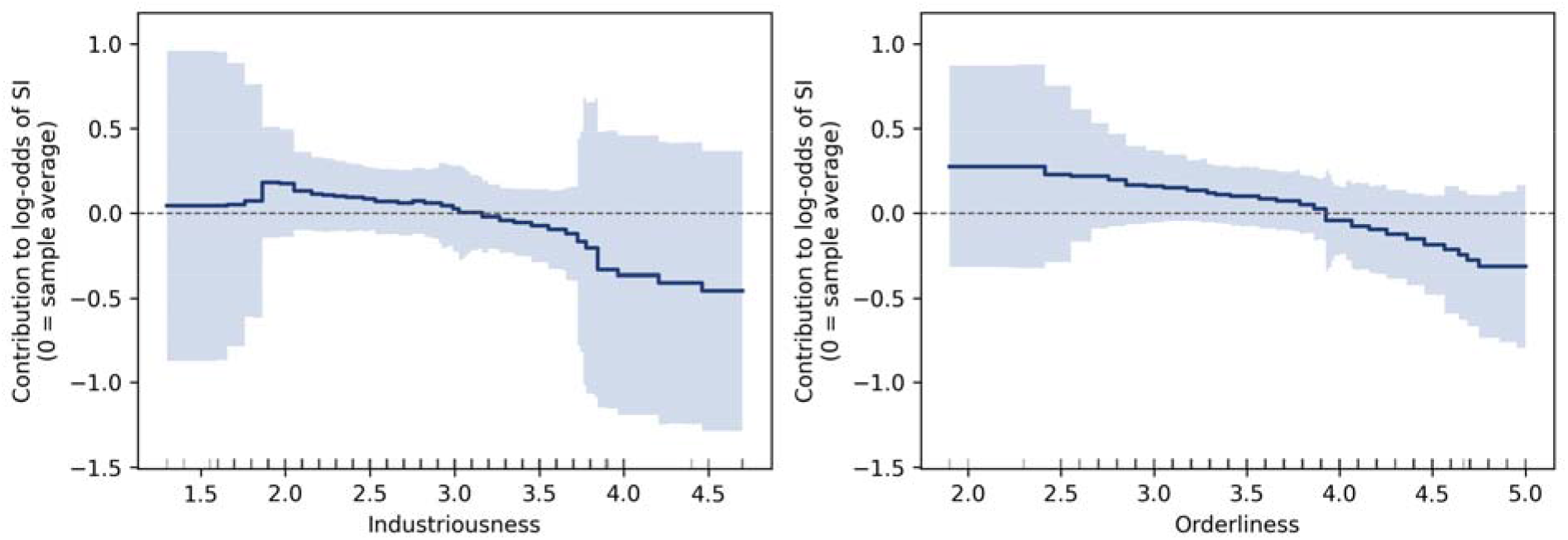
**Shape Functions for the Conscientiousness Aspects** *Note.* Contributions to the log-odds of suicidal ideation (0 = sample average) when industriousness and orderliness replace the conscientiousness domain score in the additive model. Bands show resampling stability (Section S3); ticks mark observed scores.

**Figure S3.**
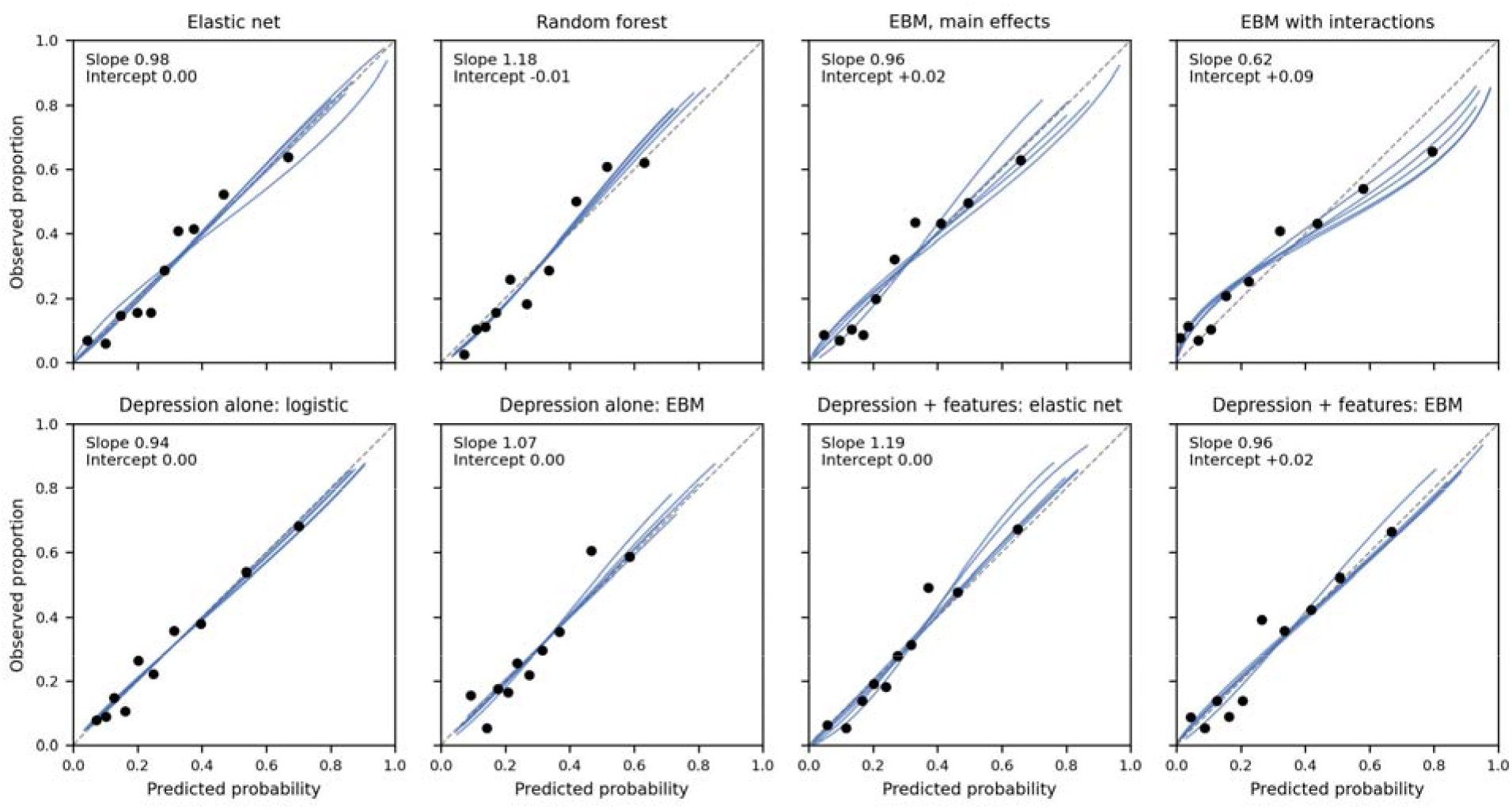
**Calibration of Out-of-Fold Predictions** *Note.* Points are observed proportions within deciles of predicted risk, pooling the five repeats of cross-validation; lines are the logistic recalibration curve of each repeat; the dashed line marks perfect calibration. Slope and intercept are the calibration slope and calibration-in-the-large, averaged across repeats. EBM = explainable boosting machine.

## S6. TRIPOD+AI Reporting Checklist

Table S10 maps the TRIPOD+AI items (Collins et al., 2024) to the manuscript. Item topics are paraphrased.

**Table S10.** TRIPOD+AI Checklist Mapping.

*TRIPOD+AI Checklist Mapping*
| Item | Topic (paraphrased) | Where reported | Note or action |
| --- | --- | --- | --- |
| *Title and abstract* |  |  |  |
| 1 | Title identifies model development, population, and outcome | Title page |  |
| 2 | Abstract (see TRIPOD+AI for Abstracts) | Abstract |  |
| *Introduction* |  |  |  |
| 3a | Health care context and rationale | Introduction, paragraphs 1–3 |  |
| 3b | Target population, intended purpose, and users | Introduction, final paragraph;<br>Discussion, Clinical Implications | Model is descriptive, not intended for deployment |
| 3c | Known health inequalities | Discussion, Limitations and Future Directions | Sample composition noted; not otherwise addressed |
| 4 | Objectives | Introduction, final paragraph |  |
| *Methods* |  |  |  |
| 5a | Data sources and representativeness | Methods, Participants |  |
| 5b | Dates of data collection | Methods, Participants | Enrollment since 2020; data extracted May 2026 |
| 6a | Setting and number of centres | Methods, Participants | Single specialty clinic |
| 6b | Eligibility criteria | Methods, Participants |  |
| 6c | Treatments received | Methods, Measures; Table 1 | Medication status as a covariate |
| 7 | Data preparation and quality checking | Methods, Analytic Plan |  |
| 8a | Outcome definition, timing, and rationale | Methods, Measures |  |
| 8b | Qualifications of outcome assessors | Not applicable | Self-report |
| 8c | Blinding of outcome assessment | Not applicable | Self-report, cross-sectional |
| 9a | Choice and pre-selection of predictors | Introduction; Methods, Features and models |  |
| 9b | Definition and measurement of predictors | Methods, Measures |  |
| 9c | Qualifications of predictor assessors | Not applicable | Self-report |
| 10 | Sample size justification | Methods, Participants; Discussion, Limitations and Future Directions | Sample set by registry availability; 66 events for 15 features (4.4 per feature) |
| 11 | Missing data | Methods, Analytic Plan | *k*-nearest-neighbor imputation within training folds; complete-case check of the in-sample models (Table S8) |
| 12a | Use of data for development and evaluation | Methods, Validation | Nested cross-validation |
| 12b | Handling of predictors | Methods, Analytic Plan |  |
| 12c | Model type, building steps, tuning, internal validation | Methods; Table S1 |  |
| 12d | Heterogeneity across clusters | Not applicable | Single site |

OCD, DEPRESSION, AND SUICIDAL IDEATION
| Item | Topic (paraphrased) | Where reported | Note or action |
| --- | --- | --- | --- |
| 12e | Performance measures and model comparisons | Methods, Validation | Corrected resampled CIs for mean performance and for differences |
| 12f | Model updating | Not applicable |  |
| 12g | How predictions were calculated | Not applicable | Development study with internal validation only |
| 13 | Class imbalance | Methods, Features and models | No reweighting; PR-AUC tuning criterion |
| 14 | Fairness | Discussion, Limitations and Future Directions | No fairness methods applied; stated in Limitations |
| 15 | Model output and classification thresholds | Methods, Validation | Youden threshold from training folds |
| 16 | Differences between development and evaluation data | Not applicable | Internal validation only |
| 17 | Ethical approval and consent | Methods, Participants |  |
| *Open science* |  |  |  |
| 18a | Funding | Title page | No funding received; stated in the author note |
| 18b | Conflicts of interest | Title page | CP's disclosures listed; BAZ and EFM have none |
| 18c | Protocol | Introduction, final paragraph; Methods, Software and reporting | No protocol prepared; stated in Methods |
| 18d | Registration | Methods, Software and reporting | Not preregistered; stated in the text |
| 18e | Data sharing | Methods, Software and reporting | Available from the corresponding author on reasonable request |
| 18f | Code sharing | Methods, Software and reporting | Analysis code in a public repository (link in Methods) |
| 19 | Patient and public involvement | Methods, Software and reporting | No patient or public involvement; stated in Methods |
| *Results* |  |  |  |
| 20a | Participant flow and outcome events | Methods, Participants; Results | 236 records; 231 analyzed; 66 with SI |
| 20b | Participant characteristics and missing data | Results; Table 1 |  |
| 20c | Comparison with development data | Not applicable | No external evaluation |
| 21 | Participants and events in each analysis | Methods, Validation | Outer training folds 184–185 participants (52–53 events); test folds 46–47 (13–14 events) |
| 22 | Full model specification | Methods, Software and reporting | Descriptive models; the shared code refits them from the data (available on request) |
| 23a | Performance with confidence intervals, including subgroups | Table 2; Table 3; Supplementary Figure S3 | Subgroup performance not assessed; stated in Limitations |
| 23b | Heterogeneity in performance across clusters | Not applicable |  |
| 24 | Model updating results | Not applicable |  |
| *Discussion* |  |  |  |
| 25 | Overall interpretation | Discussion |  |

OCD, DEPRESSION, AND SUICIDAL IDEATION
| Item | Topic (paraphrased) | Where reported | Note or action |
| --- | --- | --- | --- |
| 26 | Limitations | Discussion, Limitations and Future Directions |  |
| 27a | Handling poor quality or unavailable inputs | Not applicable | Not intended for implementation |
| 27b | User interaction and expertise | Not applicable | Not intended for implementation |
| 27c | Next steps for applicability | Discussion, Limitations and Future Directions |  |

*Note.* Item numbers follow Collins et al. (2024). PR-AUC = area under the precision-recall curve; SI = suicidal ideation.

